# Efficient blockLASSO for Polygenic Scores with Applications to All of Us and UK Biobank

**DOI:** 10.1101/2024.06.25.24309482

**Authors:** Timothy G. Raben, Louis Lello, Erik Widen, Stephen D.H. Hsu

**Affiliations:** Department of Physics and Astronomy, Michigan State University; Genomic Prediction, Inc., North Brunswick, NJ

## Abstract

We develop a “block” LASSO (blockLASSO) method for training polygenic scores (PGS) and demonstrate its use in All of Us (AoU) and the UK Biobank (UKB). BlockLASSO utilizes the approximate block diagonal structure (due to chromosomal partition of the genome) of linkage disequilibrium (LD). LASSO optimization is performed chromosome by chromosome, which reduces computational complexity by orders of magnitude. The resulting predictors for each chromosome are combined using simple re-weighting techniques. We demonstrate that blockLASSO is generally as effective for training PGS as (global) LASSO and other approaches. This is shown for 11 different phenotypes, in two different biobanks, and across 5 different ancestry groups (African, American, East Asian, European, and South Asian). The block approach works for a wide variety of phenotypes. In the past, it has been shown that some phenotypes are more/less polygenic than others. Using sparse algorithms, an accurate PGS can be trained for type 1 diabetes (T1D) using 100 single nucleotide variants (SNVs). On the other extreme, a PGS for body mass index (BMI) would need more than 10k SNVs. blockLasso produces similar PGS for phenotypes while training with just a fraction of the variants per block. For example, within AoU (using only genetic information) block PGS for T1D (1,500 cases/113,297 controls) reaches an AUC of 0.63_*±*0.02_ and for BMI (102,949 samples) a correlation of 0.21_*±*0.01_. This is compared to a traditional global LASSO approach which finds for T1D an AUC 0.65_*±*0.03_ and BMI a correlation 0.19_*±*0.03_. Similar results are shown for a total of 11 phenotypes in both AoU and the UKB and applied to all 5 ancestry groups as defined via an Admixture analysis. In all cases the contribution from common covariates – age, sex assigned at birth, and principal components – are removed before training. This new block approach is more computationally efficient and scalable than global machine learning approaches. Genetic matrices are typically stored as memory mapped instances, but loading a million SNVs for a million participants can require 8TB of memory. Running a LASSO algorithm requires holding in memory at least two matrices this size. This requirement is so large that even large high performance computing clusters cannot perform these calculations. To circumvent this issue, most current analyses use subsets: e.g., taking a representative sample of participants and filtering SNVs via pruning and thresholding. High-end LASSO training uses ∼ 500 GB of memory (e.g., ∼ 400k samples and ∼ 50k SNVs) and takes 12-24 hours to complete. In contrast, the block approach typically uses ∼ 200× (2 orders of magnitude) less memory and runs in ∼ 500× less time.

## 1 Introduction

Polygenic scores (PGS) are becoming important tools for understanding genetic architecture[1–3], identifying potential genetic risk of disease[4, 5], and now have clinical applications[6]. PGS are traditionally built in one of two ways: (1) starting from single marker regression and adding in additional genomic information or (2) training algorithms executed directly on a subset of the genome.

The first approach principally relies on the results of single marker regression, or genome wide association studies (GWAS). The GWAS results are then re-weighted using linkage disequilibrium (LD) structure (i.e., the correlation structure of SNPs), functional information, fine mapping, meta-analyses, or ancestry specific effects. This approach has the advantage that GWAS is computationally efficient (it can be run completely in parallel) and that parts of the additional information can be computed separately. Examples of this include PRS-cs[7, 8] and LDpred[9–11] which uses as inputs the results of a GWAS and the LD information, and then re-weights the SNPs using a sparse continuous shrinkage prior. This LD information can be computed a single time for a population and used in the creation of many predictors. The disadvantage of this approach is that the GWAS and LD matrices represent approximations of the genome level data and that information can be lost in this process.

The second approach relies on applying machine learning algorithms – penalized frequentist/Bayesian regressions, neural networks, decision trees, etc. [12–14] – directly on genome level data. The advantage here is that the algorithms train directly on genomes and don’t have to approximate any structure. The disadvantage is that this requires loading large genetic matrices into computer memory. For example, loading a matrix that includes 50,000 SNPs for 500,000 people at single precision requires roughly 800 gigabytes (GB) of memory.

Over the past decade there have been efforts to speed up LASSO computations by using “safe” [15] and “strong” [16] screening rules. Screening refers to identifying features that will remain with zero weight at successive LASSO hyperparameter steps, thus reducing the effective dimensionality of the problem at that step. Several works have shown possible improved computational efficiency [17–19]. For example, the *Batch Screening Iterative Lasso* (BASIL)[20] uses the “strong” rules and found a ∼ 20% faster computation time to other LASSO solutions. However, for some genomic applications, previous work that used the “safe” rules in a custom Julia implementation [21, 22] found similar results and similar computation times to a python implementation using scikit-learn without the “safe” rules [23–25]. It has also been shown that the improvements from the implementation of these screening rules depend on how they are implemented [26].

Here we utilize the underlying biological structure to greatly reduce computational time and resource requirements by computing an *approximate* LASSO solution. The main idea is that over large distances (e.g., from one chromosome to another) SNVs are uncorrelated. In other words, the correlation structure is block diagonal as in **Figure 1**. From this assumption we can run LASSO (or another algorithm) on each block independently and then perform a re-weighting to find the relative importance of each block. While the *exact* LASSO solution is not recovered, the majority of the predictive power appears to remain and the computation times are ∼ 500x faster than a traditional calculation.

**Figure 1:**
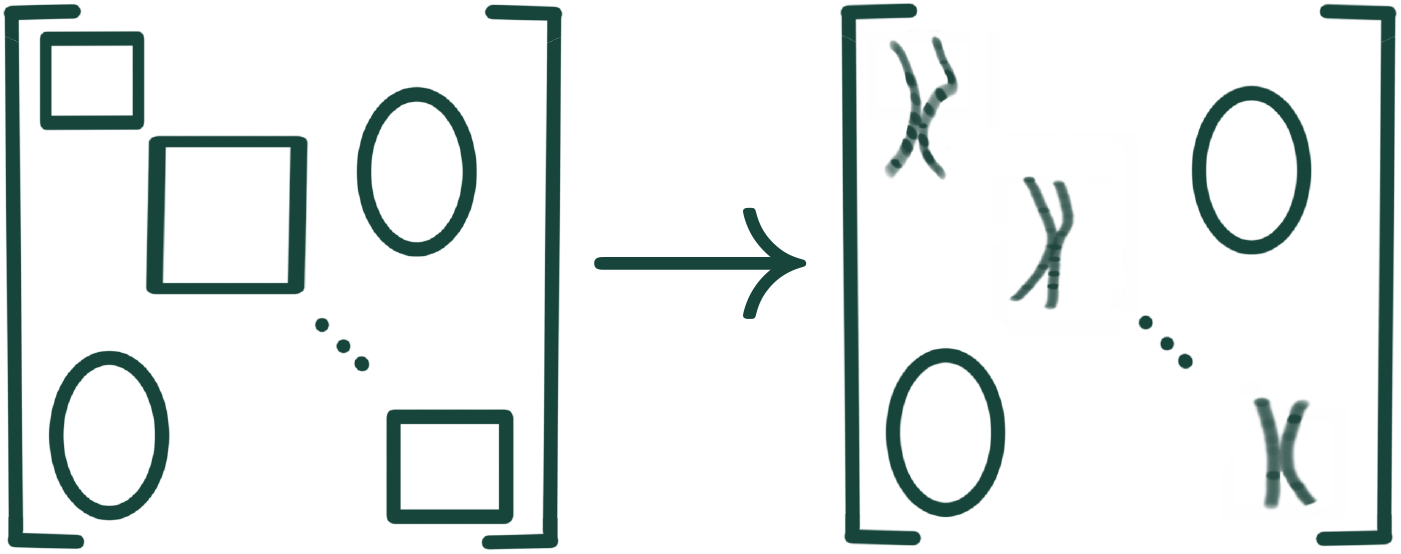
(Left) a generic block diagonal matrix. (Right) block diagonal matrices resulting from negligible correlations across (as opposed to within) chromosomes.

## 2 Results

The main results can be seen in **Table 1** and a subset are displayed in **Figure 2**, while the full set of results are collected in the Supplementary Information. Results are presented in terms of Area Under Receiver Operator Curve (AUC) for case control phenotypes and phenotype to PGS correlation (corr.) for quantitative phenotypes. All uncertainties are estimates of the standard deviation of the true distribution, i.e. are a reflection of the width of the entire distribution. *Global LASSO* refers to the standard LASSO algorithm run with 50k features spread throughout the autosome. *BlockLASSO* refers to small LASSOs run chromosome by chromosome and then stitched together as described in Section 3. **Table 1** shows the results for the block LASSO vs Global LASSO for training in All of Us (AoU), the full UK Biobank (UKB), and the UKB with training/testing sets reduced to the sizes found in AoU (UKB^*\**^), i.e., to give a more direct comparison. The global UKB^*\**^ column is computed from the Monte Carlo bounds set by interpolating between training sizes[25]. Table cells highlighted in 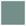 indicate that the global LASSO and blockLASSO agree (within uncertainty) in AoU – this is true for 4 case control conditions and 2 continuous phenotypes. For the other two continuous phenotypes, marked in 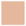, the overall signal loss from using the block approach is less than 20%.

**Table 1:**
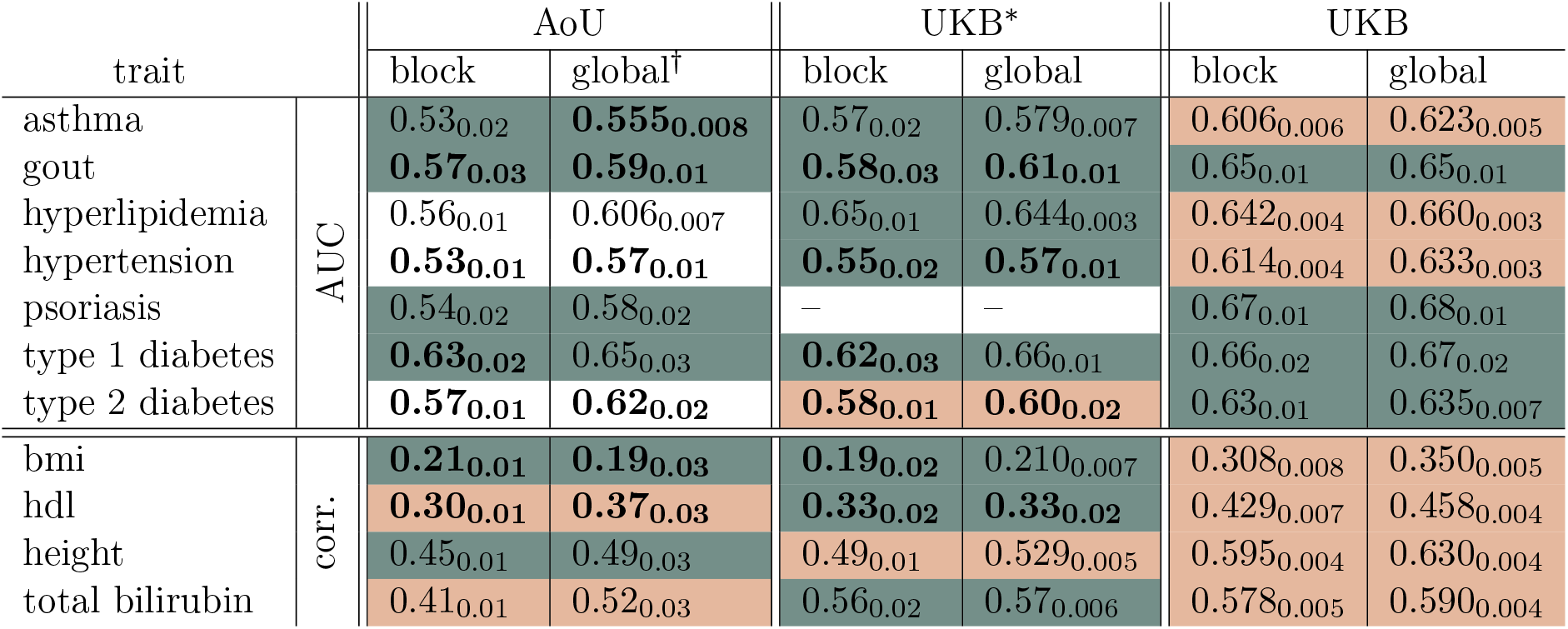
Summary of main PGS metrics for results in All of Us (AoU), the UK Biobank trained with sets matching the size of those found in AoU (UKB^*\**^), and for the UK Biobank using the maximum possible training sizes (UKB). For one trait, psoriasis, there are actually more cases in AoU than in the UKB so the UKB^*\**^ computation is left blank. All predictors are trained and tested on European populations. Blocks colored 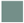 indicate that the block and traditional results agree within uncertainty. Blocks colored 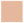 indicate that the block and traditional results disagree by less than 20%. Finally, **bold text** indicates that the results between AoU and UKB (either block or traditional) are in agreement within uncertainty.

| trait |  | AoU |  | UKB* |  | UKB |  |
| --- | --- | --- | --- | --- | --- | --- | --- |
|  |  | block | global† | block | global | block | global |
| asthma | AUC | 0.53<sub>0.02</sub> | **0.555<sub>0.008</sub>** | 0.57<sub>0.02</sub> | 0.579<sub>0.007</sub> | 0.606<sub>0.006</sub> | 0.623<sub>0.005</sub> |
| gout |  | **0.57<sub>0.03</sub>** | **0.59<sub>0.01</sub>** | **0.58<sub>0.03</sub>** | **0.61<sub>0.01</sub>** | 0.65<sub>0.01</sub> | 0.65<sub>0.01</sub> |
| hyperlipidemia |  | 0.56<sub>0.01</sub> | 0.606<sub>0.007</sub> | 0.65<sub>0.01</sub> | 0.644<sub>0.003</sub> | 0.642<sub>0.004</sub> | 0.660<sub>0.003</sub> |
| hypertension |  | **0.53<sub>0.01</sub>** | **0.57<sub>0.01</sub>** | **0.55<sub>0.02</sub>** | **0.57<sub>0.01</sub>** | 0.614<sub>0.004</sub> | 0.633<sub>0.003</sub> |
| psoriasis |  | 0.54<sub>0.02</sub> | 0.58<sub>0.02</sub> | — | — | 0.67<sub>0.01</sub> | 0.68<sub>0.01</sub> |
| type 1 diabetes |  | **0.63<sub>0.02</sub>** | 0.65<sub>0.03</sub> | **0.62<sub>0.03</sub>** | 0.66<sub>0.01</sub> | 0.66<sub>0.02</sub> | 0.67<sub>0.02</sub> |
| type 2 diabetes |  | **0.57<sub>0.01</sub>** | **0.62<sub>0.02</sub>** | **0.58<sub>0.01</sub>** | **0.60<sub>0.02</sub>** | 0.63<sub>0.01</sub> | 0.635<sub>0.007</sub> |
| bmi | corr. | **0.21<sub>0.01</sub>** | **0.19<sub>0.03</sub>** | **0.19<sub>0.02</sub>** | 0.210<sub>0.007</sub> | 0.308<sub>0.008</sub> | 0.350<sub>0.005</sub> |
| hdl |  | **0.30<sub>0.01</sub>** | **0.37<sub>0.03</sub>** | **0.33<sub>0.02</sub>** | **0.33<sub>0.02</sub>** | 0.429<sub>0.007</sub> | 0.458<sub>0.004</sub> |
| height |  | 0.45<sub>0.01</sub> | 0.49<sub>0.03</sub> | 0.49<sub>0.01</sub> | 0.529<sub>0.005</sub> | 0.595<sub>0.004</sub> | 0.630<sub>0.004</sub> |
| total bilirubin |  | 0.41<sub>0.01</sub> | 0.52<sub>0.03</sub> | 0.56<sub>0.02</sub> | 0.57<sub>0.006</sub> | 0.578<sub>0.005</sub> | 0.590<sub>0.004</sub> |

**Figure 2:**
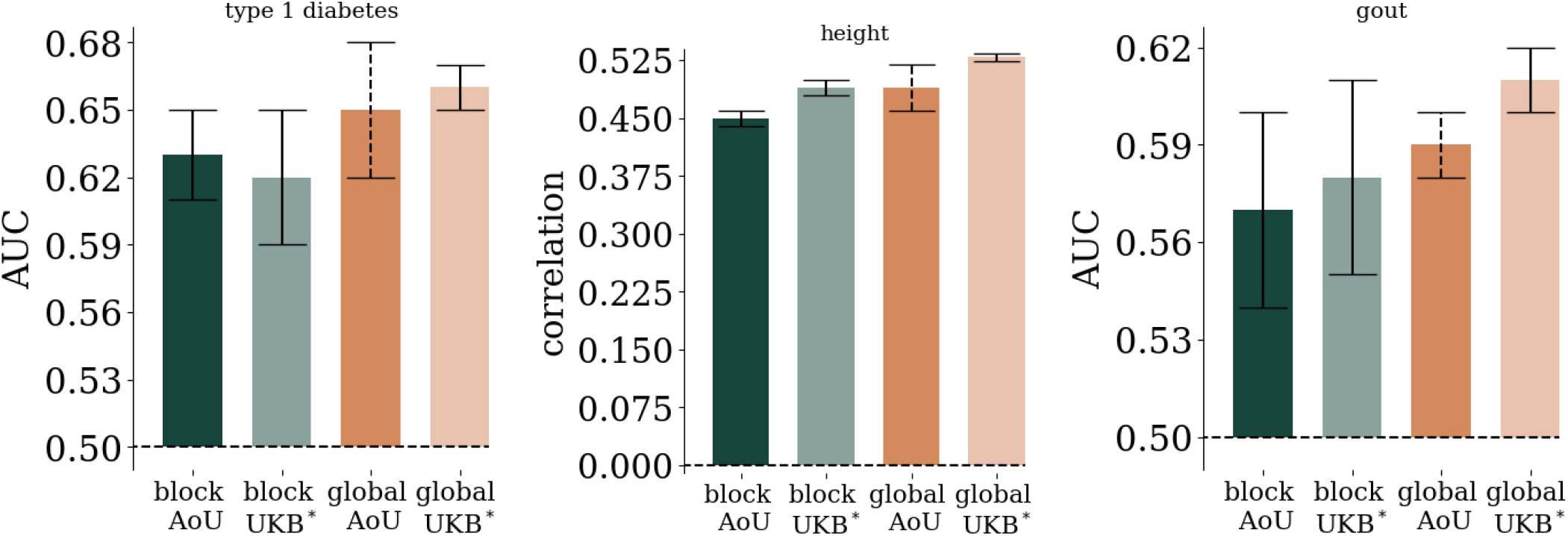
Comparison of block vs global LASSO in AoU and UKB^*\**^ (UKB reduced to training/testing sets comparable to AoU). We see that for many conditions both the block and global results are in agreement in both biobanks *and* the results are in agreement between biobanks. Uncertainties reflect one standard deviation computed from 5-fold cross-validation and computing AUC/correlation with finite sample sizes. For the “global AoU” measurement only one training fold was run so the uncertainty is the larger of the finite size effect, or the corresponding uncertainty found in the UKB.

For the reduced UK Biobank training (UKB^*\**^) we see that 5 case-control and 3 continuous phenotypes are in agreement between block and global LASSO. The remaining two phenotypes have a signal fall-off of less than 20%. Psoriasis was omitted as it is the one phenotype where there are more cases in AoU than in the UKB. In principal we could limit the number of cases on the AoU side, but this would be a different calculation than the others presented. In addition we see 9 different measurements are labeled in bold indicating that the results between AoU and UKB^*\**^ are consistent within uncertainty. There are known phenotyping errors within AoU. For example, there are BMI measurements that are ∼ 10^6^ kg*/*m^2^. Although these clearly unrealistic outlying measurements are filtered before training, there is still the possibility of residual phenotyping errors. Despite these possible phenotyping errors and using different SNP sets, the bolded results indicate that a similar level of polygenic prediction was found for 9 measurements between the two different biobanks.

Finally we see that for maximal training within the UKB 4 phenotypes agree within uncertainty between block and global LASSO and 6 phenotypes have less than a 20% signal loss. Training with the full UKB uses much larger training and testing sets which is reflected in the smaller uncertainties.

Traditional global LASSO is limited by computation time, memory requirements, and cost, all three of which can limit the number of features (SNVs) and samples (individuals) used to train models. In previous research [21, 22, 24, 25, 27], a typical global LASSO was run on ∼ 50k SNPs and ∼ 400k people. These computations required between 500-700 GB of RAM and would complete after 8-24 hours of computational time. The computations were mostly done on 28 core Intel(R) Xeon(R) CPU E5-2680 v4 (2.40 GHz) processors. Some older computations were done on 20 core Intel(R) Xeon(R) CPU E5-2670 v2 (2.50 GHz) processors.

As seen in **Figure 3** the blockLASSO approach is much faster and uses much less memory than the global LASSO. Using the full UKB we trained blockLASSO predictors using different numbers of SNPs per chromosome. There is an average increase in speed of a factor of 4.7_*±*0.8_ × 10^2^, i.e. over 2 orders of magnitude.

**Figure 3:**
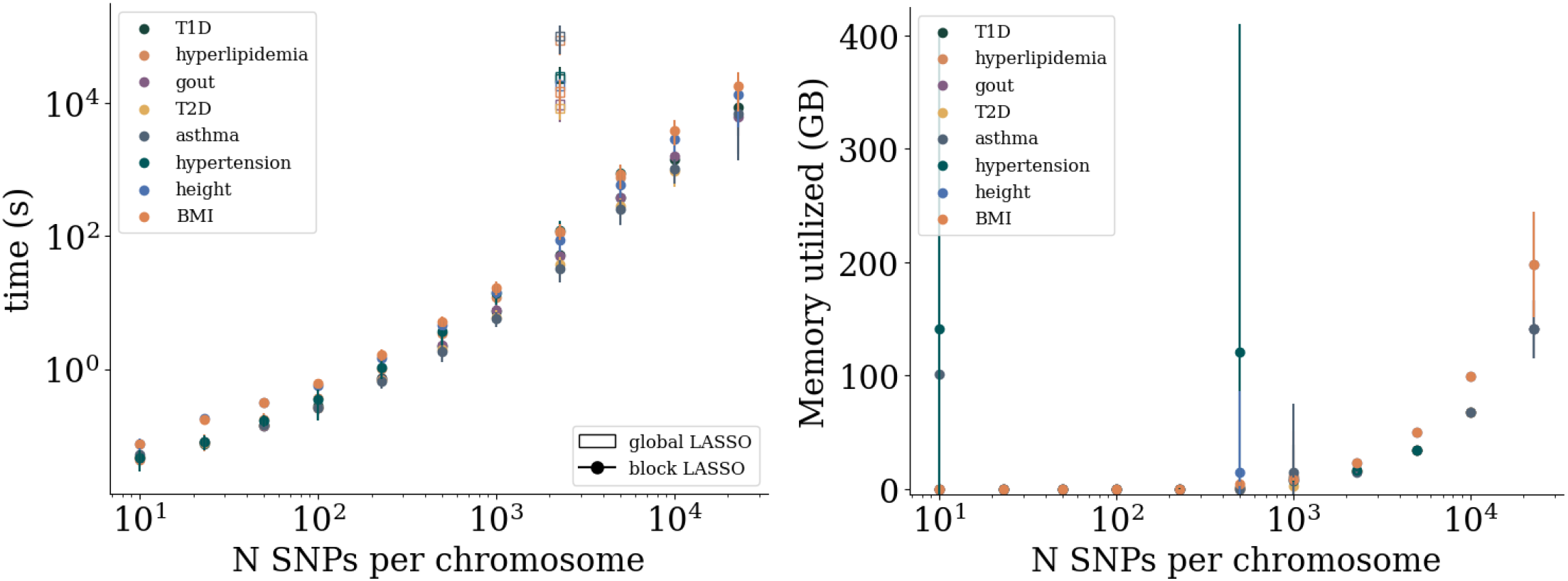
Computational details for the block LASSO in the UKB. Left: computation time (seconds) as a function of number of SNVs per chromosome. Error bars are standard deviation over runs. blockLASSO in the specific case of 2273 SNVs per autosomal chromosome is compared to the traditional global LASSO using 50,000 SNVs (or SNPs) and corresponds to an average 4.7_*±*0.8_ × 10^2^ times increase in computation time. A typical global LASSO using 50,000 SNVs typically requires 300-700 GB of memory. Right: RAM used per block LASSO SNPs per chromosome. Error bars are standard deviations over runs. For most phenotypes and SNP sizes the error bars are very small. For SNVs per chromosome below 10^3^ the larger error bars reflect runs that did not converge and contribute trivially to the overall performance, i.e., these were runs where the coordinate descent algorithm failed to converge and the resulting AUC(correlation) was ∼ 0.5(0) respectively.

In **Figure 4** we see in the top panel the effective variance fraction explained by variants across the autosome. As described in section 3.5, this approximate variance per location is computed by summing over one dimension of the feature correlation matrix. Error bars reflect the uncertainty over cross-validation folds, but only one fold is used for the global AoU results. Binning, and adding error bars in quadrature, of nearby positions is done to make the plot human readable. In the bottom panel, these contributions are summed to show the variance fraction explained per chromosome. Again, the error bars come from using multiple cross-validation folds. By comparing the block LASSO to the traditional LASSO we see that the block method recovers the same important regions with similar weights. Also analogously, by comparing the results from the UKB and AoU we see similar variance fractions explained in similar regions.

**Figure 4:**
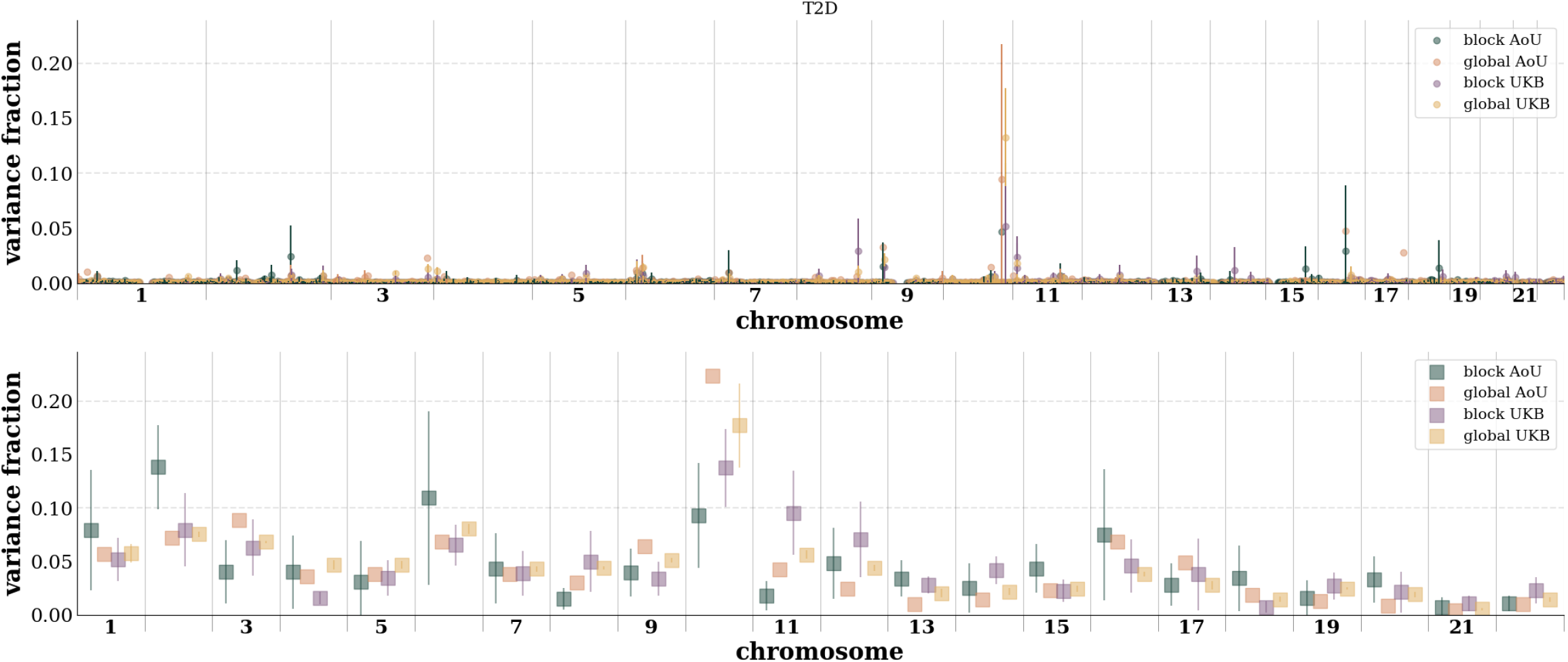
Comparison of block vs global LASSO binned variance per base pair position (top) and per chromosome (bottom) in AoU and the UKB for type 2 diabetes. This approximate variance is identified by computing the PGS covariance matrix and summing over rows as detailed in section 3.5. Error bars are standard deviations over cross-validation folds and then propagated in quadrature as nearby contributions are binned (top) or summed over the entire chromosome (bottom). For global LASSO in AoU only a single CV fold was computed. Similar plots for the other phenotypes can be found in Supplementary Information.

## 3 Methods

### 3.1 Phenotypes

The phenotypes used are a combination of self-report/survey data, ICD9/10 code diagnoses, and laboratory measurements. Exact combinations that make up each phenotype in both AoU and UKB can be found in the Supplementary Information. Additionally, we include the number of samples for each phenotype used in training. In general we do not use exclusion criteria, i.e. we have no criteria that would exclude a documented case or control (e.g., age of onset or minimum age). The only exception is for *psoriasis in AoU* where we found a less inclusive definition of cases and controls allowed for better results. While there are accurate PGS for psoriasis (e.g., [28–30]), it is also known that there are various types of psoriasis that can have separate genetic signatures [31–33]. For this phenotype alone, we use a set of exclusion criteria for both cases and controls.

Some pre-filtering is done on continuous phenotypes to exclude values that may reflect a rare genetic condition (e.g., dwarfism), possibly reflective of extreme lifestyle choices (extreme obesity), and to exclude obvious data errors. In AoU we use the following cuts on continuous phenotypes: BMI <60 kg*/*m^2^ to filter implausible values, 144 cm < height < 205 cm to filter out possibly distinct genetics such as dwarfism and Marfan syndrome, total bilirubin < 2.5 mg*/*dl to filter unrealistic values, and hdl <200 mg*/*dl to filter unrealistic values. In the UKB we require that measurements be positive values and that height >144 cm.

### 3.2 Populations

Within the UKB we largely relied on self-reported “Ethnic background” responses to identify ancestry. We used the following mapping from UKB fields: Asian or Asian British, Indian, Pakistani, Bangladeshi, Any other Asian background–South Asian (SAS); Black or Black British, Caribbean, African, Any other Black background–African (AFR); Chinese–East Asian (EAS); and White, British, Irish, Any other white background–European (EUR).

For both UKB and AoU we also computed genetic ancestry via ADMIXTURE [34]. This was used to identify a generalized American or Hispanic group in UKB and also general identification in AoU. We implemented Admixture on “1000 Genomes” [35] with 5 groupings – this generated an ancestry fraction for each individual for each of the 5 groups and we found that the ancestry fractions corresponded with their respective superpopulations (EUR, AFR, AMR, SAS, EAS). These results were then applied to the UK Biobank where it was found that the ancestry fractions largely aligned with the 4 self-reported categories. Individuals with at least 0.35 ancestry fraction in the fifth grouping, which corresponded to AMR in 1000 Genomes, were labeled as American (AMR) and excluded from the other groupings. Within AoU we relied exclusively on the admixture approach but compared ancestry fractions against survey results for consistency. Again 5 groupings were used with ancestry fraction cuts of 0.7 for AFR, 0.3 for AMR, 0.8 for EAS, 0.9 for EUR, and 0.6 for SAS.

Within the UKB we identify genetic siblings to be used as a testing set (not used in training). Genetic siblings can be assumed to have more similar environmental backgrounds than random pairs. This leads to slightly reduced PGS accuracy as discussed in [23, 36]. The method for identifying siblings is described in [23] in supplemental section C.

### 3.3 Polygenic scores

This project demonstrates an efficient approach to building PGS that is applied block by block on the genome. To this end we are interested in *only the genetic component* of the PGS. Future work will be aimed at combining this approach with other effects (environmental, gene × environment interactions, etc.) to generate maximal possible prediction accuracy. To train these genetic-only predictors we take an approach to try to be as conservative as possible. The process can be briefly summarized as follows: (1) separate samples into training, model selection (validation), and testing sets; (2) regress phenotype on covariates and build residual phenotype; (3) identify candidate SNPs (4) perform block by block LASSO (5) re-weight relative block LASSO results (6) evaluate on completely withheld testing sets.

(1) For all phenotypes we split the samples into training, model-selection/re-weighting, and testing sets. In both biobanks we used the EUR cohort for training as it had the largest sample sizes. We remove a small subset of the EUR group and the entire AFR, AMR, EAS, and SAS groups to be used for final testing sets. In the UKB the EUR testing set is a group of sibling pairs as described above. After removing the testing sets we assemble training and model-selection sets using 5-fold cross-validation. Exact set sizes are given in the Supplementary Information.

Next we build residual phenotypes as step (2) using the training sets. For a case-control phenotype we regress the raw phenotype on covariates – age, sex, and principal components (top 16 in AoU and top 20 in UKB). For continuous phenotypes we do sex-specific z-scoring so that we can combine the sex assorted data into a single, much larger training set. We then use linear regression on age and principal components. For all phenotypes we then subtract off the effect of these covariates to make a residual phenotype. Building residual phenotypes allow us to give *conservative* estimates of the genetic effects since we first assume that the covariates have a *maximal* impact and then we train the genetics on just the remaining information. Additionally, because PRS have not be extensively trained in AoU we did global LASSO training for case-control phenotypes on the raw phenotype allowing us to be conservative about our statement of agreement between block and global results.

Step (3) involves identifying candidate features (SNVs). For both global and block LASSO approaches we perform a GWAS using the training set and rank SNVs by p-values across the entire autosome or block (chromosome) respectively. This GWAS is used purely to rank SNVs and *none* of the other GWAS information (e.g., p-values, odds ratios, weights) is retained or used. In previous research involving case-control phenotypes in the UKB it was found that rank ordering results from a GWAS on the *raw phenotype* (e.g., [22, 23]) lead to similar results as rank ordering results from a GWAS on *residual* phenotype (e.g., [25]). For the block approach, *within the UKB*, this remained true. However, when we moved to AoU, we found, only for the block approach, that selecting features based on a GWAS of the raw phenotype under-performed selecting features based on a GWAS of the residual phenotype. This is likely a result of the AoU biobank being more diverse both environmentally and ancestrally causing these covariates to have a much larger effect. When selecting the top features we used a cut off to include only SNVs with allele frequency *>* 0.001 in the training population to avoid spurious associations.

For the global approach it has been previously shown that the top 50k SNVs are more than sufficient to build a PGS [21, 22]. It was shown that the resultant sparse PGS included SNVs all throughout the top 50k SNVs, but that increasing the total number of SNVs (e.g., to 100k[21]) did not noticeably improve results, even for the least sparse predictors. For the block approach, the ideal number of features is not a priori known. It will not necessarily be the case that the set of SNVs from the 50k top ranked SNVs *across the entire autosome* will be the same set as the collection of the top 2,273 SNVs on each autosomal chromosome. To this end we tested different block sizes: {10; 23; 50; 100; 227; 500; 1,000; 2,273; 5,000; 10,000; 22,727} SNVs. The performance of training with different block sizes within the UKB can be seen in **Figure 5** and in the Supplementary Information. We see that performance tends to plateau near 2,273 SNVs per chromosome (50,006 SNVs over the entire autosome) which is roughly equivalent to the size of the global LASSO training. This plateau behavior generally persists when tested in other ancestry groups.

**Figure 5:**
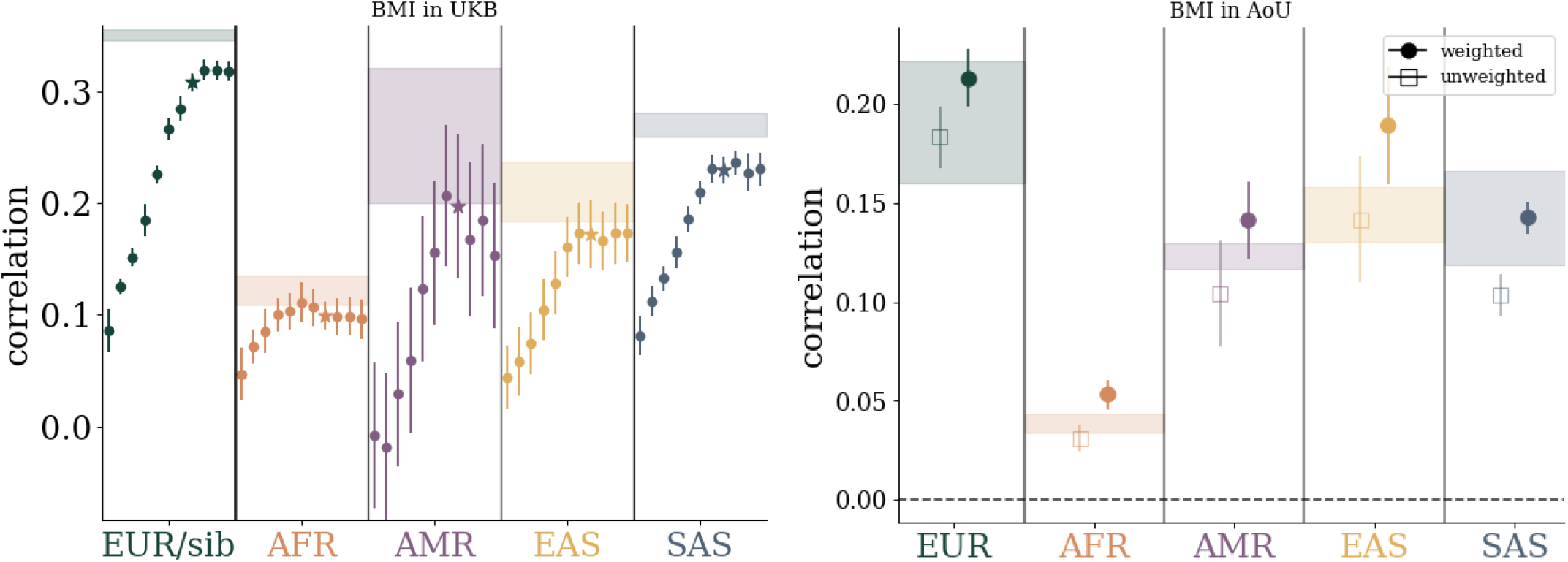
Left: performance as a function of training SNV size in UKB and applied to different ancestry groups. Within each ancestry group, dots correspond to training with {10, 23, 50, 100, 227, 500, 1000, 2273, 5000, 10000, 22727} SNVs per chromosome from left to right respectively. The starred data points correspond to 2273 SNVs per chromosome wich is roughly equivalent to 50k SNVs across the autosome. Right: performance before and after the re-weighting step of the blockLASSO. While re-weighting is trained within the EUR group, the effect of re-weighting improves prediction accuracy across all tested ancestry groupings.

After feature selection, step (4) involves running the LASSO algorithm. We used Scikit-Learn with settings specified in Supplementary Information. Model selection is done by selecting the maximal performance in the validation/model-selection set.

Step (5) is only for the blockLASSO. Using the validation/model-selection set we now do a linear regression of block scores against the raw phenotype. In principle, the re-weighting does not need to be done with linear regression, but because there are so few features in the current example (i.e., 22 chromosomes), testing with more advanced algorithms yielded the same results as linear regression. An example of the effect of this re-weighting, for all 5 ancestry groups, can be seen on the right side of **Figure 5**.

Finally, in step (6) the model (i.e., individual SNV weights *and* block weights) is applied to the testing sets, which have been witheld from all training steps.

### 3.4 PGS performance uncertainties

When using the block LASSO approach, the reported uncertainty is a standard deviation (i.e., reflects the width of the overall possible distribution) and has contributions from 5-fold cross-validation and from computing metrics with finite sample sizes. These two effects are added in quadrature. The only exception is the global LASSO in AoU which is both slow and costly. For this alone we only use a single global LASSO run to compute metrics and the uncertainties are reported as the larger of either the uncertainty from the finite sample size computation of the metric *or* the uncertainty in the UKB with the same training size. Specific definitions for the finite sample size contributions are given in the Supplementary Information.

### 3.5 PGS variance distributions

In order to estimate the localized contribution to the variance of the PGS we compute an approximate covariance per SNV (feature). To do this we compute the covariance between each feature and sum the contribution from all correlated features. I.e., we build a covariance matrix of features and sum over columns. The sum of the partially summed covariance is the total variance of the PGS. The explicit mathematical definition is given in the Supplementary Information. This approximate variance per locus is a generalized version of “single SNP variance” defined in [27, 37]. When training with *n* samples (e.g., people) and *m* total features (e.g., all SNVs from all blocks), the covariance between features is an *m* × *m* matrix. In **Figure 4**, and in the Supplementary Information, we see the result of summing over one axis in the covariance matrix. To make the plot more human-readable we filter out variants accounting for less than 0.01% of the total variance and by binning nearby variants within 1 Mbp. After cutting and binning we attribute this partial sum to the effective location of the binned variants (a weighted average over variant location). Error bars are computed by averaging over cross-validation folds.

## 4 Conclusion

There are many successful methods for training PGS. Generally, compared to methods that rely on summary statistics or genome wide associations, methods that train directly on genotypes and involve variant/nucleotide level data can produce equivalent (or better) results with much less data (e.g., human height prediction using ∼ 450k individuals in [21] vs ∼ 5.4 million individuals[38]). Additionally, many of the limitations associated with PGS in general are actually limitations of *GWAS-based* PGS (e.g., [39]). There is an added interest in *sparse* methods, i.e., algorithms that perform feature selection, allowing inferences regarding genetic architecture and pleiotropy. Currently, most commonly used sparse methods perform similarly, with the simple least absolute shrinkage and selection operator (LASSO) regularly performing among the best methods[25]. Sparse methods are often advantageous as performing feature selection can reduce the computational demand. However, training directly on genotypes – even after pre-filtering SNVs – is still very demanding and typically requires hundreds of gigabytes of memory and tens of hours of run time on a computing cluster. Here we present an improved method which makes use of block diagonal structure in SNP correlations associated with independent chromosomes (or other large regions). For a PGS that trains with 50k SNPs and 450k samples, this produces a ∼ 75x reduction in required memory and compute resources.

LASSO based predictors have previously been trained in one biobank and tested in a separate biobank with only minimal loss in performance (e.g., [22]). It is encouraging here that we see training and testing a polygenic predictor is largely independent of dataset (i.e. AoU vs UKB) – given equal data size. This suggests confounders – such as intake, genotyping platform/procedure, societal differences, etc. – have diminutive effect on the interpretations of these LASSO based PGS trained directly on genotypes.

We hope that the novel method demonstrated in this work can be improved upon. While screening rules – i.e., rules which aid in feature selection – have only resulted in modest improvements for the global LASSO approaches, it is possible that incorporating screening will further speed up block approaches. Some PGS models have been improved via ancestry specific tagging and SNV selection[40, 41], functional information[42–45], and more careful definitions of phenotypes[46–48]. It should be possible to integrate all of these approaches with the new method presented here.

Finally we mention that initial research has shown the implementation of PGS in the clinic can lead to significantly better outcomes for some diagnoses[49]. There is also a large literature highlighting potential benefits for identifying those at high risk of particular diseases, aiding in early detection, reducing total cost of care. This is all evidence for PGS as a generally useful tool in clinical practice[22, 50–54]. However, there are still outstanding practical [55–59] and ethical [60, 61] challenges related to the widespread adoption of PGS to the practice of clinical medicine.

## Data Availability

All data produced in this present study are either available online at https://github.com/MSU-Hsu-Lab/blockLASSO or available upon reasonable request to the authors.

## 5 Acknowledgements

Analysis using UK Biobank data used computational resources hosted by the Michigan State University High-Performance Computing Center. The authors acknowledge acquisition of data sets via UK Biobank Main Application 15326. UK Biobank linked health data acknowledgement: Data from the UK Biobank includes data provided by patients and collected by the National Health Service (NHS) England as part of their care and support. UK Biobank data also includes data assets made available by National Safe Haven as part of the Data and Connectivity National Core Study, led by Health Data Research UK in partnership with the Office for National Statistics and funded by UK Research and Innovation (research which commenced between 1st October 2020–31st March 2021 grant ref MC_PC_20029; 1st April 2021–30th September 2022 grant ref MC_PC_20058).

Work with AoU data was performed using the All of Us Researcher Workbench under the workspace “Scalable and efficient polygenic scores in diverse populations”. We are grateful and acknowledge the contributions of the *All of Us* participants who make this project possible and the work of the National Institutes of Health’s *All of Us* Research Program for making this data available.

## 7 Software

Analysis, data management, and figure generation used Python 3. As detailed below, specific packages and functions can be found in the associated code examples. ADMIXTURE version 1.3.0 was used in genetic ancestry analysis.

## 8 Data availability

Block predictors from both the UKB and AoU and code examples can be found in the Hsu group GitHub: https://github.com/MSU-Hsu-Lab/blockLASSO. Exact code used to generate the AoU results can be found in the “Scalable and efficient polygenic scores in diverse populations” workspace in the AoU “controlled tier” data access tier.

## 9 Abbreviations

AFR: African
AMR: American
AoU: All of Us
AUC: area under receiver operator curve
BASIL: batch screening iterative LASSO
BMI: body mass index
EAS: East Asian
EUR: European
GB: gigabyte
GWAS: genome wide association studies
hdl: high-density lipoprotein
LASSO: least absolute shrinkage and selection operator
LD: linkage disequilibrium
Mbp: megabase pair
PGS: polygenic score
SAS: South American
SNP: single nucleotide polymorphism
SNV: single nucleotide variant
T1D: type 1 diabetes
T2D: type 2 diabetes
UKB: UK Biobank

## References

1. Masip, G. et al. The genetic architecture of the association between eating behaviors and obesity: combining genetic twin modeling and polygenic risk scores. The American Journal of Clinical Nutrition 112, 956–966 (2020) (cit. on p. 1).

2. Trinder, M., Vikulova, D., Pimstone, S., Mancini, G. J. & Brunham, L. R. Polygenic architecture and cardiovascular risk of familial combined hyperlipidemia. Atherosclerosis 340, 35–43 (2022) (cit. on p. 1).

3. Wang, Y. et al. Polygenic prediction across populations is influenced by ancestry, genetic architecture, and methodology. Cell Genomics 3 (2023) (cit. on p. 1).

4. Busby, G. B. et al. Ancestry-specific polygenic risk scores are risk enhancers for clinical cardiovascular disease assessments. Nature Communications 14, 7105 (2023) (cit. on p. 1).

5. Baliakas, P. et al. Integrating a Polygenic Risk Score into a clinical setting would impact risk predictions in familial breast cancer. Journal of Medical Genetics 61, 150–154 (2024) (cit. on p. 1).

6. Yang, X., Kar, S., Antoniou, A. C. & Pharoah, P. D. Polygenic scores in cancer. Nature reviews Cancer 23, 619–630 (2023) (cit. on p. 1).

7. Ge, T., Chen, C.-Y., Ni, Y., Feng, Y.-C.A. & Smoller, J. W. Polygenic prediction via Bayesian regression and continuous shrinkage priors. Nature communications 10, 1–10 (2019) (cit. on p. 2).

8. Ruan, Y. et al. Improving Polygenic Prediction in Ancestrally Diverse Populations. medRxiv, 2020.12.27.20248738. 10.1101/2020.12.27.20248738v2%0A https://www.medrxiv.org/content/10.1101/2020.12.27.20248738v2.abstract (2021) (cit. on p. 2).

9. Vilhjálmsson, B. et al. Modeling linkage disequilibrium increases accuracy of polygenic risk scores. The american journal of human genetics 97, 576–592 (2015) (cit. on p. 2).

10. Márquez-Luna, C. et al. LDpred-funct: incorporating functional priors improves polygenic prediction accuracy in UK Biobank and 23andMe data sets. BioRxiv, 375337 (2020) (cit. on p. 2).

11. Privé, F., Arbel, J. & Vilhjálmsson, B. J. LDpred2: better, faster, stronger. Bioinformatics 36, 5424–5431 (2020) (cit. on p. 2).

12. Perez, P. & de los Campos, G. Genome-Wide Regression and Prediction with the BGLR Statistical Package. Genetics 198, 483–495. 10.1534/genetics.114.164442 (July 2014) (cit. on p. 2).

13. Yokoyama, J. S. et al. Decision tree analysis of genetic risk for clinically heterogeneous Alzheimer’s disease. BMC neurology 15, 1–11 (2015) (cit. on p. 2).

14. Badré, A., Zhang, L., Muchero, W., Reynolds, J. C. & Pan, C. Deep neural network improves the estimation of polygenic risk scores for breast cancer. Journal of Human Genetics 66, 359–369 (2021) (cit. on p. 2).

15. Ghaoui, L. E., Viallon, V. & Rabbani, R. Safe Feature Elimination in Sparse Supervised Learning. Pacific Journal of Optimization 8, 667–698 (4 Jan. 2012) (cit. on p. 2).

16. Tibshirani, R. et al. Strong rules for discarding predictors in lasso-type problems. Journal of the Royal Statistical Society Series B: Statistical Methodology 74, 245–266 (2012) (cit. on p. 2).

17. Liu, J., Zhao, Z., Wang, J. & Ye, J. Safe screening with variational inequalities and its application to lasso. arXiv preprint 1307.7577 (2013) (cit. on p. 2).

18. Liu, J., Zhao, Z., Wang, J. & Ye, J. Safe Screening with Variational Inequalities and Its Application to Lasso in Proceedings of The 31st International Conference on Machine Learning (eds Xing, E.P. & Jebara, T.) 32 (JMLR Workshop and Conference Proceedings, Jan. 2014), 289–297 (cit. on p. 2).

19. Malti, A. & Herzet, C. Safe screening tests for LASSO based on firmly non-expansiveness in 2016 IEEE International Conference on Acoustics, Speech and Signal Processing (ICASSP) (IEEE, Mar. 2016). 10.1109/icassp.2016.7472575 (cit. on p. 2).

20. Qian, J. et al. A fast and scalable framework for large-scale and ultrahigh-dimensional sparse regression with application to the UK biobank. PLoS Genetics. issn: 15537404 (2020) (cit. on p. 2).

21. Lello, L. et al. Accurate genomic prediction of human height. Genetics 210. [PMC6216598], 477–497 (2018) (cit. on pp. 2, 4, 7, 8).

22. Lello, L., Raben, T. G., Yong, S. Y., Tellier, L. C. & Hsu, S. D. H. Genomic prediction of 16 complex disease risks including heart attack, diabetes, breast and prostate cancer. Sci Rep 9. [PMC6814833], 1–16 (2019) (cit. on pp. 2, 4, 7, 9).

23. Lello, L., Raben, T. G. & Hsu, S. D. H. Sibling validation of polygenic risk scores and complex trait prediction. Scientific Reports 10. [PMC7411027], 13190. 10.1038/s41598-020-69927-7 (2020) (cit. on pp. 2, 6, 7).

24. Widen, E., Raben, T. G., Lello, L. & Hsu, S. D. H. Machine Learning Prediction of Biomarkers from SNPs and of Disease Risk from Biomarkers in the UK Biobank. Genes 12. issn: 2073-4425. https://www.mdpi.com/2073-4425/12/7/991 (2021) (cit. on pp. 2, 4).

25. Raben, T. G., Lello, L., Widen, E. & Hsu, S. D. Biobank-scale methods and projections for sparse polygenic prediction from machine learning. Scientific Reports 13, 11662 (2023) (cit. on pp. 2–4, 7, 9).

26. Fercoq, O., Gramfort, A. & Salmon, J. Mind the duality gap: safer rules for the Lasso. ArXiv e-prints. 1505.03410 [stat.ML] (May 2015) (cit. on p. 2).

27. Yong, S. Y., Raben, T. G., Lello, L. & Hsu, S. D. Genetic Architecture of Complex Traits and Disease Risk Predictors. Scientific Reports 10. [PMC7374622] (2020) (cit. on pp. 4, 8).

28. Yin, X. et al. A weighted polygenic risk score using 14 known susceptibility variants to estimate risk and age onset of psoriasis in Han Chinese. PloS one 10, e0125369 (2015) (cit. on p. 6).

29. Shen, M. et al. Associations of combined lifestyle and genetic risks with incident psoriasis: A prospective cohort study among UK Biobank participants of European ancestry. Journal of the American Academy of Dermatology 87, 343–350 (2022) (cit. on p. 6).

30. Yang, J.-S. et al. Genome-wide association study and polygenic risk scores predict psoriasis and its shared phenotypes in Taiwan. Molecular Medicine Reports 30, 1–22 (2024) (cit. on p. 6).

31. Dand, N. et al. Psoriasis and genetics (cit. on p. 6).

32. Raharja, A., Mahil, S. K. & Barker, J. N. Psoriasis: a brief overview. Clinical Medicine 21, 170 (2021) (cit. on p. 6).

33. Soomro, M. et al. Comparative genetic analysis of psoriatic arthritis and psoriasis for the discovery of genetic risk factors and risk prediction modeling. Arthritis & rheumatology 74, 1535–1543 (2022) (cit. on p. 6).

34. Alexander, D. H., Novembre, J. & Lange, K. Fast model-based estimation of ancestry in unrelated individuals. Genome research 19, 1655–1664 (2009) (cit. on p. 6).

35. Auton, A. et al. A global reference for human genetic variation. Nature 526, 68–74. 10.1038/nature15393 (2015) (cit. on p. 6).

36. Lello, L., Hsu, M., Widen, E. & Raben, T. G. Sibling variation in polygenic traits and DNA recombination mapping with UK Biobank and IVF family data. Scientific Reports 13, 376. 10.1038/s41598-023-27561-z (2023) (cit. on p. 6).

37. Raben, T. G., Lello, L., Widen, E. & Hsu, S. D. Biobank-scale methods and projections for sparse polygenic prediction from machine learning. medRxiv. eprint: https://www.medrxiv.org/content/early/2023/03/08/2023.03.06.23286870.full.pdf. https://www.medrxiv.org/content/early/2023/03/08/2023.03.06.23286870 (2023) (cit. on p. 8).

38. Yengo, L. et al. A saturated map of common genetic variants associated with human height. Nature 610, 704–712 (2022) (cit. on p. 8).

39. Yair, S. & Coop, G. Population differentiation of polygenic score predictions under stabilizing selection. Philosophical Transactions of the Royal Society B 377, 20200416 (2022) (cit. on p. 8).

40. Amariuta, T. et al. Improving the trans-ancestry portability of polygenic risk scores by prioritizing variants in predicted cell-type-specific regulatory elements. Nature Genetics 52, 1346–1354. issn: 15461718. 10.1038/s41588-020-00740-8 (2020) (cit. on p. 9).

41. Shi, H. et al. Population-specific causal disease effect sizes in functionally important regions impacted by selection. Nature Communications 12. issn: 20411723. 10.1038/s41467-021-21286-1 (2021) (cit. on p. 9).

42. Kichaev, G. et al. Integrating functional data to prioritize causal variants in statistical fine-mapping studies. PLoS genetics 10, e1004722 (2014) (cit. on p. 9).

43. Hu, Y. et al. Joint modeling of genetically correlated diseases and functional annotations increases accuracy of polygenic risk prediction. PLoS genetics 13, e1006836 (2017) (cit. on p. 9).

44. Marquez-Luna, C. et al. Modeling functional enrichment improves polygenic prediction accuracy in UK Biobank and 23andMe data sets. bioRxiv. eprint: https://www.biorxiv.org/content/early/2018/07/24/375337.full.pdf. https://www.biorxiv.org/content/early/2018/07/24/375337 (2018) (cit. on p. 9).

45. Weissbrod, O. et al. Functionally informed fine-mapping and polygenic localization of complex trait heritability. Nature Genetics 52. issn: 15461718. 10.1038/s41588-020-00735-5 (2020) (cit. on p. 9).

46. Carroll, R. J., Bastarache, L. & Denny, J. C. R PheWAS: data analysis and plotting tools for phenome-wide association studies in the R environment. Bioinformatics 30, 2375–2376 (2014) (cit. on p. 9).

47. Leader, J. B. et al. Contrasting association results between existing PheWAS phenotype definition methods and five validated electronic phenotypes in AMIA Annual Symposium Proceedings 2015 (2015), 824 (cit. on p. 9).

48. Wu, P. et al. Mapping ICD-10 and ICD-10-CM codes to phecodes: workflow development and initial evaluation. JMIR medical informatics 7, e14325 (2019) (cit. on p. 9).

49. Samani, N. J. et al. Polygenic risk score adds to a clinical risk score in the prediction of cardiovascular disease in a clinical setting. European Heart Journal, ehae342 (2024) (cit. on p. 9).

50. Iribarren, C. et al. Clinical utility of multimarker genetic risk scores for prediction of incident coronary heart disease: a cohort study among over 51 000 individuals of European ancestry. Circulation: Cardiovascular Genetics 9, 531–540 (2016) (cit. on p. 9).

51. Inouye, M. et al. Genomic Risk Prediction of Coronary Artery Disease in 480,000 Adults: Implications for Primary Prevention. Journal of the American College of Cardiology 72, 1883 –1893. issn: 0735-1097. http://www.sciencedirect.com/science/article/pii/S0735109718369493 (2018) (cit. on p. 9).

52. O’Sullivan, J. W. et al. Polygenic risk scores for cardiovascular disease: a scientific statement from the American Heart Association. Circulation 146, e93–e118 (2022) (cit. on p. 9).

53. Lu, X. et al. A polygenic risk score improves risk stratification of coronary artery disease: a large-scale prospective Chinese cohort study. European heart journal 43, 1702–1711 (2022) (cit. on p. 9).

54. Marston, N. A. et al. Predictive Utility of a Coronary Artery Disease Polygenic Risk Score in Primary Prevention. JAMA Cardiology 8, 130–137. issn: 2380-6583. eprint: https://jamanetwork.com/journals/jamacardiology/articlepdf/2800101/jamacardiology\_marston\_2022\_oi\_220077\_1675366811.13075.pdf. 10.1001/jamacardio.2022.4466 (Feb. 2023) (cit. on p. 9).

55. Wray, N. R. et al. From basic science to clinical application of polygenic risk scores: a primer. JAMA psychiatry 78, 101–109 (2021) (cit. on p. 9).

56. Hao, L. et al. Development of a clinical polygenic risk score assay and reporting workflow. Nature medicine 28, 1006–1013 (2022) (cit. on p. 9).

57. Klarin, D. & Natarajan, P. Clinical utility of polygenic risk scores for coronary artery disease. Nature Reviews Cardiology 19, 291–301 (2022) (cit. on p. 9).

58. Koch, S., Schmidtke, J., Krawczak, M. & Caliebe, A. Clinical utility of polygenic risk scores: a critical 2023 appraisal. Journal of Community Genetics 14, 471–487 (2023) (cit. on p. 9).

59. Xiang, R. et al. Recent advances in polygenic scores: translation, equitability, methods and FAIR tools. Genome Medicine 16, 33 (2024) (cit. on p. 9).

60. Lewis, A. C. & Green, R. C. Polygenic risk scores in the clinic: new perspectives needed on familiar ethical issues. Genome Medicine 13, 1–10 (2021) (cit. on p. 9).

61. Abu-El-Haija, A. et al. The clinical application of polygenic risk scores: A points to consider statement of the American College of Medical Genetics and Genomics (ACMG). Genetics in Medicine 25, 100803 (2023) (cit. on p. 9).

