## Supplemental material for "Efficient blockLASSO for Polygenic Scores with Applications to All of Us and UK Biobank"

### Efficient Block Lasso for building polygenic scores with applications to the UK Biobank and All of Us

June 25, 2024

**3 PGS metrics plots** **11**

**4 Variance plots** **13**

**5 Training and re-weighting** **19**

**6 LASSO validation paths** **25**

### 1 Data

Phenotypes from the UK Biobank (UKB) are based upon the November 20th, 2023 release of UKB data. Case control codes are made from `Non-cancer illness code, self-reported`, `Diagnoses - ICD9`, and `Diagnoses - ICD10` codes. Exact sample counts can be found in **Table 1**. The following definitions are used in the UKB:

**type 2 diabetes** non-cancer codes: `1223`
ICD9: `25000, 25002, 25010, 25012, 25020, 25022, 25030, 25032, 25040, 25042, 25050, 25052, 25060, 25062, 25070, 25072, 25080, 25082, 25090, 25092`
ICD10: `E11, E110, E111, E112, E113, E114, E115, E116, E117, E118, E119`

**asthma** non-cancer codes: `1111`
ICD9: `49300, 49309 ,49310 ,49319 ,49390 ,49399`
ICD10: `J450,J451,J458`

**gout** noncancer: `1466`
ICD9: `2740, 2741, 2748, 2749, 7120`
ICD10: `M1000, M1001, M1002, M1003, M1004, M1005, M1006, M1007, M1008, M1009, E790`

**psoriasis** noncancer: `1453`
ICD9: `6961, 6962, 6968`
ICD10: `L400, L401, L404, L405, L408, L409, L413, L414,L415, L418, L419, M0900, M0901, M0902, M0903,M0904, M0905, M0906, M0907, M0908, M0909`

**hyperlipidemia** field ID `30690` (total cholesterol) with value $\geq 6.21$ as case

**type 1 diabetes** noncancer: `1222`
ICD10: `E100, E101, E102, E103, E104, E105, E106, E107, E108, E109, O240`

**hypertension** noncancer: `1065,1072,1073`
ICD9: `4010, 4011, 4019, 4050, 4051, 4059, 4160, 6420, 6423, 6429`
ICD10: `I10`

**height** field ID: `50`

**bmi** field ID: `21001`

**total bilirubin** field ID: `30840`

**hdl** field ID: `30760`

Phenotypes from AoU were constructed using the AoU workbench. Exact sample sizes can be found in **Table 2**. Using keyword searchers, most case control phenotypes all included information from "All Surveys" data and continuous phenotypes can be found in the "physical measurements" survey data. Additionally, the following concepts were used in combination with the survey results.

**type 2 diabetes** Standard Concepts: Acidosis due to type 2 diabetes mellitus; Angina associated with type 2 diabetes mellitus; Arthropathy due to type 2 diabetes mellitus; Autonomic neuropathy due to type 2 diabetes mellitus; Cataract due to diabetes mellitus type 2; Chronic kidney disease due to type 2 diabetes mellitus; Chronic kidney disease stage 2 due to type 2 diabetes mellitus; Chronic kidney disease stage 3 due to type 2 diabetes mellitus; Chronic kidney disease stage 4 due to type 2 diabetes mellitus; Chronic kidney disease stage 5 due to type 2 diabetes mellitus; Coronary artery disease due to type 2 diabetes mellitus; Dermopathy due to type 2 diabetes mellitus; Diabetes

| phenotype | training | | model selection | | testing | |
|---|---|---|---|---|---|---|
| | cases | controls | cases | controls | cases | controls |
| asthma | 48,775 | 151,225 | 1,250 | 1,250 | 4,589 | 35,499 |
| gout | 7,188 | 192,812 | 1,250 | 1,250 | 795 | 39,293 |
| hyperlipidemia | 125,414 | 272,412 | 1,250 | 1,250 | 12,477 | 27,611 |
| hypertension | 167,219 | 249,072 | 1,250 | 1,250 | 16,039 | 24,049 |
| psoriasis | 7,740 | 192,260 | 1,250 | 1,250 | 906 | 39,182 |
| type 1 diabetes | 2,629 | 197,371 | 1,250 | 1,250 | 368 | 39,720 |
| type 2 diabetes | 29,588 | 170,412 | 1,250 | 1,250 | 2,833 | 37,255 |
| phenotype | training | model selection | testing | | | |
| bmi | 415,082 | 2,500 | 40,088 | | | |
| hdl | 364,809 | 2,500 | 40,088 | | | |
| height | 414,960 | 2,500 | 40,088 | | | |
| total bilirubin | 396,219 | 2,500 | 40,088 | | | |

**Table 1:** Phenotype sample sizes in UKB. For case control conditions with less than 50k cases, the controls were limited to keep a total of 200k samples to speed up training.

| phenotype | training | | model selection | | testing | |
|---|---|---|---|---|---|---|
| | cases | controls | cases | controls | cases | controls |
| asthma | 20,866 | 90,931 | 1,000 | 1,000 | 1,000 | 1,000 |
| gout | 2,477 | 112,320 | 250 | 250 | 250 | 250 |
| hyperlipidemia | 51,013 | 60,784 | 1,000 | 1,000 | 1,000 | 1,000 |
| hypertension | 46,371 | 65,426 | 1,000 | 1,000 | 1,000 | 1,000 |
| psoriasis | 2,588 | 84,269 | 500 | 500 | 500 | 500 |
| type 1 diabetes | 1,500 | 113,297 | 250 | 250 | 250 | 250 |
| type 2 diabetes | 15,522 | 96,275 | 1,000 | 1,000 | 1,000 | 1,000 |
| phenotype | training | model selection | testing | | | |
| bmi | 102,949 | 5,000 | 5,000 | | | |
| hdl | 44,665 | 5,000 | 5,000 | | | |
| height | 103,202 | 5,000 | 5,000 | | | |
| total bilirubin | 56,776 | 5,000 | 5,000 | | | |

**Table 2:** Phenotype sample sizes in AoU

mellitus type 2 without retinopathy; Disorder due to type 2 diabetes mellitus; Disorder due to well controlled type 2 diabetes mellitus; Disorder of eye due to type 2 diabetes mellitus; Disorder of nervous system due to type 2 diabetes mellitus; Dyslipidemia due to type 2 diabetes mellitus; End stage renal disease on dialysis due to type 2 diabetes mellitus; Erectile dysfunction due to type 2 diabetes mellitus; Foot ulcer due to type 2 diabetes mellitus; Gangrene due to type 2 diabetes mellitus; Gastroparesis due to type 2 diabetes mellitus; Hyperglycemia due to type 2 diabetes mellitus; Hyperosmolar coma due to type 2 diabetes mellitus; Hyperosmolar non-ketotic state due to type 2 diabetes mellitus; Hypoglycemia due to type 2 diabetes mellitus; Hypoglycemic coma due to type 2 diabetes mellitus; Insulin treated type 2 diabetes mellitus; Ketoacidosis due to type 2 diabetes mellitus; Ketoacidotic coma due to type 2 diabetes mellitus; Macular edema and retinopathy due to type 2 diabetes mellitus; Macular edema due to type 2 diabetes mellitus; Microalbuminuria due to type 2 diabetes mellitus; Mild nonproliferative retinopathy due to type 2 diabetes mellitus; Mixed hyperlipidemia due to type 2 diabetes mellitus; Moderate nonproliferative retinopathy due to type 2 diabetes mellitus; Mononeuropathy due to type 2 diabetes mellitus; Multiple complications due to type 2 diabetes mellitus; Neuropathic arthropathy due to type 2 diabetes mellitus; Neuropathy due to type 2 diabetes mellitus; Nonproliferative retinopathy due to type 2 diabetes mellitus; Peripheral circulatory disorder due to type 2 diabetes mellitus; Peripheral neuropathy due to type 2 diabetes mellitus; Peripheral sensory neuropathy due to type 2 diabetes mellitus; Polyneuropathy due to type 2 diabetes mellitus; Pre-existing type 2 diabetes mellitus; Pre-existing type 2 diabetes mellitus in pregnancy; Pregnancy and type 2 diabetes mellitus; Proliferative retinopathy due to type 2 diabetes mellitus; Proteinuria due to type 2 diabetes mellitus; Renal disorder due to type 2 diabetes mellitus; Retinal edema due to type 2 diabetes mellitus; Retinopathy due to type 2 diabetes mellitus; Traction detachment of retina due to type 2 diabetes mellitus; Type 2 diabetes mellitus; Type 2 diabetes mellitus controlled by diet; Type 2 diabetes mellitus in nonobese; Type 2 diabetes mellitus in obese; Type 2 diabetes mellitus well controlled; Type 2 diabetes mellitus with peripheral angiopathy; Type 2 diabetes mellitus with ulcer; Type 2 diabetes mellitus without complication; Ulcer of heel due to type 2 diabetes mellitus; Ulcer of left foot due to type 2 diabetes mellitus; Ulcer of lower limb due to type 2 diabetes mellitus; Ulcer of right foot due to type 2 diabetes mellitus; Ulcer of toe due to type 2 diabetes mellitus

Source Concepts: Pre-existing type 2 diabetes mellitus, in childbirth; Pre-existing type 2 diabetes mellitus, in pregnancy; Pre-existing type 2 diabetes mellitus, in pregnancy, childbirth and the puerperium; Pre-existing type 2 diabetes mellitus, in pregnancy, first trimester; Pre-existing type 2 diabetes mellitus, in pregnancy, second trimester; Pre-existing type 2 diabetes mellitus, in pregnancy, third trimester; Pre-existing type 2 diabetes mellitus, in pregnancy, unspecified trimester; Pre-existing type 2 diabetes mellitus, in the puerperium; Type 2 diabetes mellitus; Type 2 diabetes mellitus with circulatory complications; Type 2 diabetes mellitus with diabetic amyotrophy; Type 2 diabetes mellitus with diabetic arthropathy; Type 2 diabetes mellitus with diabetic autonomic (poly)neuropathy; Type 2 diabetes mellitus with diabetic cataract; Type 2 diabetes mellitus with diabetic chronic kidney disease; Type 2 diabetes mellitus with diabetic dermatitis; Type 2 diabetes mellitus with diabetic macular edema, resolved following treatment; Type 2 diabetes mellitus with diabetic macular edema, resolved following treatment, bilateral; Type 2 diabetes mellitus with diabetic macular edema, resolved following treatment, left eye; Type 2 diabetes mellitus with diabetic macular edema, resolved following treatment, right eye; Type 2 diabetes mellitus with diabetic macular edema, resolved following treatment, unspecified eye; Type 2 diabetes mellitus with diabetic mononeuropathy; Type 2 diabetes mellitus with diabetic nephropathy; Type 2 diabetes mellitus with diabetic neuropathic arthropathy; Type 2 diabetes mellitus with diabetic neuropathy, unspecified; Type 2 diabetes mellitus with diabetic peripheral angiopathy with gangrene; Type 2 diabetes mellitus with diabetic peripheral angiopathy without gangrene; Type 2 diabetes mellitus with diabetic polyneuropathy; Type 2 diabetes mellitus with foot ulcer; Type 2 diabetes mellitus with hyperglycemia; Type 2 diabetes mellitus with hyperosmolarity; Type 2 diabetes mellitus with hyperosmolarity with coma; Type 2 diabetes mellitus with hyperosmolarity without nonketotic hyperglycemic-hyperosmolar coma (NKHHC); Type 2 diabetes mellitus with hypoglycemia; Type 2 diabetes mellitus with hypoglycemia with coma; Type 2 diabetes mellitus with hypoglycemia without coma; Type 2 diabetes mellitus with ketoacidosis; Type 2 diabetes mellitus with ketoacidosis with coma; Type 2 diabetes mellitus with ketoacidosis without coma; Type 2 diabetes mellitus with kidney complications; Type 2 diabetes mellitus with mild nonproliferative diabetic retinopathy; Type 2 diabetes mellitus with mild nonproliferative diabetic retinopathy with macular edema; Type 2 diabetes mellitus with mild nonproliferative diabetic retinopathy with macular edema, bilateral; Type 2 diabetes mellitus with mild nonproliferative diabetic retinopathy with macular edema, left eye; Type 2 diabetes mellitus with mild nonproliferative diabetic retinopathy with macular edema, right eye; Type 2 diabetes mellitus with mild nonproliferative diabetic retinopathy with macular edema, unspecified eye; Type 2 diabetes mellitus with mild nonproliferative diabetic retinopathy without macular edema; Type 2 diabetes mellitus with mild nonproliferative diabetic retinopathy without macular edema, bilateral; Type 2 diabetes mellitus with mild nonproliferative diabetic retinopathy without macular edema, left eye; Type 2 diabetes mellitus with mild nonproliferative diabetic retinopathy without macular edema, right eye; Type 2 diabetes mellitus with mild nonproliferative diabetic retinopathy without macular edema, unspecified eye; Type 2 diabetes mellitus with moderate nonproliferative diabetic retinopathy; Type 2 diabetes mellitus with moderate nonproliferative diabetic retinopathy with macular edema; Type 2 diabetes mellitus with moderate nonproliferative diabetic retinopathy with macular edema, bilateral; Type 2 diabetes mellitus with moderate nonproliferative diabetic retinopathy with macular edema, left eye; Type 2 diabetes mellitus with moderate nonproliferative diabetic retinopathy with macular edema, right eye; Type 2 diabetes mellitus with moderate nonproliferative diabetic retinopathy with macular edema, unspecified eye; Type 2 diabetes mellitus with moderate nonproliferative diabetic retinopathy without macular edema; Type 2 diabetes mellitus with moderate nonproliferative diabetic retinopathy without macular edema, bilateral; Type 2 diabetes mellitus with moderate nonproliferative diabetic retinopathy without macular edema, left eye; Type 2 diabetes mellitus with moderate nonproliferative diabetic retinopathy without macular edema, right eye; Type 2 diabetes mellitus with moderate nonproliferative diabetic retinopathy without macular edema, unspecified eye; Type 2 diabetes mellitus with neurological complications; Type 2 diabetes mellitus with ophthalmic complications; Type 2 diabetes mellitus with oral complications; Type 2 diabetes mellitus with other circulatory complications; Type 2 diabetes mellitus with other diabetic arthropathy; Type 2 diabetes mellitus with other diabetic kidney complication; Type 2 diabetes mellitus with other diabetic neurological complication; Type 2 diabetes mellitus with other diabetic ophthalmic complication; Type 2 diabetes mellitus with other oral complications; Type 2 diabetes mellitus with other skin complications; Type 2 diabetes mellitus with other skin ulcer; Type 2 diabetes mellitus with other specified complication; Type 2 diabetes mellitus with other specified complications; Type 2 diabetes mellitus with periodontal disease; Type 2 diabetes mellitus with proliferative diabetic retinopathy; Type 2 diabetes mellitus with proliferative diabetic retinopathy with combined traction retinal detachment and rhegmatogenous retinal detachment; Type 2 diabetes mellitus with proliferative diabetic retinopathy with combined traction retinal detachment and rhegmatogenous retinal detachment, bilateral; Type 2 diabetes mellitus with proliferative diabetic retinopathy with combined traction retinal detachment and rhegmatogenous retinal detachment, left eye; Type 2 diabetes mellitus with proliferative diabetic retinopathy with combined traction retinal detachment and rhegmatogenous retinal detachment, right eye; Type 2 diabetes mellitus with proliferative diabetic retinopathy with combined traction retinal detachment and rhegmatogenous retinal detachment, unspecified eye; Type 2 diabetes mellitus with proliferative diabetic retinopathy with macular edema; Type 2 diabetes mellitus with proliferative diabetic retinopathy with macular edema, bilateral; Type 2 diabetes mellitus with proliferative diabetic retinopathy with macular edema, left eye; Type 2 diabetes mellitus with proliferative diabetic retinopathy with macular edema, right eye; Type 2 diabetes mellitus with proliferative diabetic retinopathy with macular edema, unspecified eye; Type 2 diabetes mellitus with proliferative diabetic retinopathy with traction retinal detachment involving the macula; Type 2 diabetes mellitus with proliferative diabetic retinopathy with traction retinal detachment involving the macula, bilateral; Type 2 diabetes mellitus with proliferative diabetic retinopathy with traction retinal detachment involving the macula, left eye; Type 2 diabetes mellitus with proliferative diabetic retinopathy with traction retinal detachment involving the macula, right eye; Type

2 diabetes mellitus with proliferative diabetic retinopathy with traction retinal detachment involving the macula, unspecified eye; Type 2 diabetes mellitus with proliferative diabetic retinopathy with traction retinal detachment not involving the macula; Type 2 diabetes mellitus with proliferative diabetic retinopathy with traction retinal detachment not involving the macula, bilateral; Type 2 diabetes mellitus with proliferative diabetic retinopathy with traction retinal detachment not involving the macula, left eye; Type 2 diabetes mellitus with proliferative diabetic retinopathy with traction retinal detachment not involving the macula, right eye; Type 2 diabetes mellitus with proliferative diabetic retinopathy with traction retinal detachment not involving the macula, unspecified eye; Type 2 diabetes mellitus with proliferative diabetic retinopathy without macular edema; Type 2 diabetes mellitus with proliferative diabetic retinopathy without macular edema, bilateral; Type 2 diabetes mellitus with proliferative diabetic retinopathy without macular edema, left eye; Type 2 diabetes mellitus with proliferative diabetic retinopathy without macular edema, right eye; Type 2 diabetes mellitus with proliferative diabetic retinopathy without macular edema, unspecified eye; Type 2 diabetes mellitus with severe nonproliferative diabetic retinopathy; Type 2 diabetes mellitus with severe nonproliferative diabetic retinopathy with macular edema; Type 2 diabetes mellitus with severe nonproliferative diabetic retinopathy with macular edema, bilateral; Type 2 diabetes mellitus with severe nonproliferative diabetic retinopathy with macular edema, left eye; Type 2 diabetes mellitus with severe nonproliferative diabetic retinopathy with macular edema, right eye; Type 2 diabetes mellitus with severe nonproliferative diabetic retinopathy with macular edema, unspecified eye; Type 2 diabetes mellitus with severe nonproliferative diabetic retinopathy without macular edema; Type 2 diabetes mellitus with severe nonproliferative diabetic retinopathy without macular edema, bilateral; Type 2 diabetes mellitus with severe nonproliferative diabetic retinopathy without macular edema, left eye; Type 2 diabetes mellitus with severe nonproliferative diabetic retinopathy without macular edema, right eye; Type 2 diabetes mellitus with severe nonproliferative diabetic retinopathy without macular edema, unspecified eye; Type 2 diabetes mellitus with skin complications; Type 2 diabetes mellitus with stable proliferative diabetic retinopathy; Type 2 diabetes mellitus with stable proliferative diabetic retinopathy, bilateral; Type 2 diabetes mellitus with stable proliferative diabetic retinopathy, left eye; Type 2 diabetes mellitus with stable proliferative diabetic retinopathy, right eye; Type 2 diabetes mellitus with stable proliferative diabetic retinopathy, unspecified eye; Type 2 diabetes mellitus with unspecified complications; Type 2 diabetes mellitus with unspecified diabetic retinopathy; Type 2 diabetes mellitus with unspecified diabetic retinopathy with macular edema; Type 2 diabetes mellitus with unspecified diabetic retinopathy without macular edema; Type 2 diabetes mellitus without complications

**asthma** Standard Concept: Asthma

Source Concepts: Asthma; Asthma, unspecified; Asthma, unspecified type, unspecified; Asthma, unspecified type, with (acute) exacerbation; Asthma, unspecified type, with status asthmaticus; Chronic obstructive asthma; Chronic obstructive asthma with (acute) exacerbation; Chronic obstructive asthma with status asthmaticus; Chronic obstructive asthma, unspecified; Cough variant asthma; Cough variant asthma; Eosinophilic asthma; Extrinsic asthma; Extrinsic asthma with (acute) exacerbation; Extrinsic asthma with status asthmaticus; Extrinsic asthma, unspecified; Intrinsic asthma; Intrinsic asthma with (acute) exacerbation; Intrinsic asthma with status asthmaticus; Intrinsic asthma, unspecified; Mild intermittent asthma; Mild intermittent asthma with (acute) exacerbation; Mild intermittent asthma with status asthmaticus; Mild intermittent asthma, uncomplicated; Mild persistent asthma; Mild persistent asthma with (acute) exacerbation; Mild persistent asthma with status asthmaticus; Mild persistent asthma, uncomplicated; Moderate persistent asthma; Moderate persistent asthma with (acute) exacerbation; Moderate persistent asthma with status asthmaticus; Moderate persistent asthma, uncomplicated; Other and unspecified asthma; Other asthma; Other forms of asthma; Severe persistent asthma; Severe persistent asthma with (acute) exacerbation; Severe persistent asthma with status asthmaticus; Severe persistent asthma, uncomplicated; Unspecified asthma; Unspecified asthma with (acute) exacerbation; Unspecified asthma with status asthmaticus; Unspecified asthma, uncomplicated

**gout** Source Concepts: Gout due to renal impairment; Gout, unspecified; Idiopathic gout

**psoriasis** *cases:*

Source Concepts: Other psoriasis; Other psoriasis and similar disorders; Psoriasis; Psoriatic arthropathy

*control exclusions:*

Source Concepts: Adult-onset Still's disease; Bullous dermatoses; Bullous disorder, unspecified; Bullous disorders in diseases classified elsewhere; Chronic postrheumatic arthropathy [Jaccoud]; Congenital cutaneous mastocytosis; Congenital ichthyosis; Congenital malformation of skin, unspecified; Dermatitis factitia [artefacta]; Dermatitis herpetiformis; Dermatoglyphic anomalies; Discoid lupus erythematosus of eyelid; Discoid lupus erythematosus of eyelid; Dyschromia; Ectodermal dysplasia (anhidrotic); Epidermolysis bullosa; Erythema annulare centrifugum; Erythema in diseases classified elsewhere; Erythema intertrigo; Erythema marginatum; Erythema multiforme; Erythema nodosum; Erythematosquamous dermatosis; Erythematous condition, unspecified; Erythematous conditions; Exfoliation due to erythematous conditions according to extent of body surface involved; Exfoliative dermatitis; Factitial dermatitis; Febrile neutrophilic dermatosis [Sweet]; Felty's syndrome; Granuloma annulare; Ichthyosis congenita; Incontinentia pigmenti; Infantile (acute) (chronic) eczema; Infective dermatitis; Inflammatory polyarthropathy; Juvenile arthritis; Lennox-Gastaut syndrome; Lichen; Lichen nitidus; Lichen planopilaris; Lichen planus; Lichen simplex chronicus and prurigo; Lichen striatus; Lichenification and lichen simplex chronicus; Lupus erythematosus; Meningitis in sarcoidosis; Other autoinflammatory syndromes; Other chronic figurate erythema; Other disorders of pigmentation; Other rheumatoid arthritis with rheumatoid factor; Other specified bullous disorders; Other specified congenital anomalies of skin; Other specified congenital malformations of skin; Other specified erythematous conditions; Other specified rheumatoid arthritis; Other vasculitis limited to the skin; Parapsoriasis; Pemphigoid; Pemphigus; Pityriasis alba; Pityriasis rosea; Pityriasis rubra pilaris; Poikiloderma of Civatte; Poikiloderma vasculare atrophicans; Polyarthritis, unspecified; Prurigo; Psoriasis; Psoriasis and similar disorders; Rheumatoid arthritis and other inflammatory polyarthropathies; Rheumatoid arthritis with involvement of other organs and systems; Rheumatoid arthritis with rheumatoid factor without organ or systems involvement; Rheumatoid arthritis with rheumatoid factor, unspecified; Rheumatoid arthritis without rheumatoid factor; Rheumatoid arthritis, unspecified; Rheumatoid bursitis, ankle and foot; Rheumatoid bursitis, elbow; Rheumatoid bursitis, hand; Rheumatoid bursitis, hip; Rheumatoid bursitis, knee; Rheumatoid bursitis, multiple sites; Rheumatoid bursitis, shoulder; Rheumatoid bursitis, unspecified site; Rheumatoid bursitis, wrist; Rheumatoid heart disease with rheumatoid arthritis of ankle and foot; Rheumatoid heart disease with rheumatoid arthritis of elbow; Rheumatoid heart disease with rheumatoid arthritis of hand; Rheumatoid heart disease with rheumatoid arthritis of knee; Rheumatoid heart disease with rheumatoid arthritis of multiple sites; Rheumatoid heart disease with rheumatoid arthritis of shoulder; Rheumatoid heart disease with rheumatoid arthritis of unspecified site; Rheumatoid heart disease with rheumatoid arthritis of wrist; Rheumatoid lung disease with rheumatoid arthritis; Rheumatoid myopathy with rheumatoid arthritis of ankle and foot; Rheumatoid myopathy with rheumatoid arthritis of hand; Rheumatoid myopathy with rheumatoid arthritis of multiple sites; Rheumatoid myopathy with rheumatoid arthritis of unspecified site; Rheumatoid nodule; Rheumatoid polyneuropathy with rheumatoid arthritis of hand; Rheumatoid polyneuropathy with rheumatoid arthritis of hip; Rheumatoid polyneuropathy with rheumatoid arthritis of knee; Rheumatoid polyneuropathy with rheumatoid arthritis of multiple sites; Rheumatoid polyneuropathy with rheumatoid arthritis of shoulder; Rheumatoid polyneuropathy with rheumatoid arthritis of unspecified site; Rheumatoid polyneuropathy with rheumatoid arthritis of wrist; Rheumatoid vasculitis with rheumatoid arthritis; Rosacea; Sarcoidosis; Sarcoidosis; Seborrheic dermatitis; Seborrheic keratosis; Staphylococcal scalded skin syndrome; Subcorneal pustular dermatitis; Systemic lupus erythematosus; Systemic lupus erythematosus (SLE); Toxic erythema; Vascular disorders of skin; Vasculitis limited to the skin, unspecified; Vitiligo; Xeroderma pigmentosum

**hyperlipidemia** Standard Concept: Hypercholesterolemia

Source Concepts: Hyperlipidemia, unspecified; Mixed hyperlipidemia; Mixed hyperlipidemia; Other and unspecified hyperlipidemia; Other hyperlipidemia; Other hyperlipidemia

**type 1 diabetes** Source Concepts: Pre-existing type 1 diabetes mellitus, in childbirth; Pre-existing type 1 diabetes mellitus, in pregnancy, first trimester; Pre-existing type 1 diabetes mellitus, in pregnancy, second trimester; Pre-existing type 1 diabetes mellitus, in pregnancy, third trimester; Pre-existing type 1 diabetes mellitus, in pregnancy, unspecified trimester; Pre-existing type 1 diabetes mellitus, in the puerperium; Type 1 diabetes mellitus; Type 1 diabetes mellitus with circulatory complications; Type 1 diabetes mellitus with diabetic amyotrophy; Type 1 diabetes mellitus with diabetic arthropathy; Type 1 diabetes mellitus with diabetic autonomic (poly)neuropathy; Type 1 diabetes mellitus with diabetic cataract; Type 1 diabetes mellitus with diabetic chronic kidney disease; Type 1 diabetes mellitus with diabetic dermatitis; Type 1 diabetes mellitus with diabetic macular edema, resolved following treatment; Type 1 diabetes mellitus with diabetic macular edema, resolved following treatment, bilateral; Type 1 diabetes mellitus with diabetic macular edema, resolved following treatment, right eye; Type 1 diabetes mellitus with diabetic mononeuropathy; Type 1 diabetes mellitus with diabetic nephropathy; Type 1 diabetes mellitus with diabetic neuropathic arthropathy; Type 1 diabetes mellitus with diabetic neuropathy, unspecified; Type 1 diabetes mellitus with diabetic peripheral angiopathy with gangrene; Type 1 diabetes mellitus with diabetic peripheral angiopathy without gangrene; Type 1 diabetes mellitus with diabetic polyneuropathy; Type 1 diabetes mellitus with foot ulcer; Type 1 diabetes mellitus with hyperglycemia; Type 1 diabetes mellitus with hypoglycemia; Type 1 diabetes mellitus with hypoglycemia with coma; Type 1 diabetes mellitus with hypoglycemia without coma; Type 1 diabetes mellitus with ketoacidosis; Type 1 diabetes mellitus with ketoacidosis with coma; Type 1 diabetes mellitus with ketoacidosis without coma; Type 1 diabetes mellitus with kidney complications; Type 1 diabetes mellitus with mild nonproliferative diabetic retinopathy; Type 1 diabetes mellitus with mild nonproliferative diabetic retinopathy with macular edema; Type 1 diabetes mellitus with mild nonproliferative diabetic retinopathy with macular edema, bilateral; Type 1 diabetes mellitus with mild nonproliferative diabetic retinopathy with macular edema, left eye; Type 1 diabetes mellitus with mild nonproliferative diabetic retinopathy with macular edema, right eye; Type 1 diabetes mellitus with mild nonproliferative diabetic retinopathy with macular edema, unspecified eye; Type 1 diabetes mellitus with mild nonproliferative diabetic retinopathy without macular edema; Type 1 diabetes mellitus with mild nonproliferative diabetic retinopathy without macular edema, bilateral; Type 1 diabetes mellitus with mild nonproliferative diabetic retinopathy without macular edema, left eye; Type 1 diabetes mellitus with mild nonproliferative diabetic retinopathy without macular edema, right eye; Type 1 diabetes mellitus with mild nonproliferative diabetic retinopathy without macular edema, unspecified eye; Type 1 diabetes mellitus with moderate nonproliferative diabetic retinopathy; Type 1 diabetes mellitus with moderate nonproliferative diabetic retinopathy with macular edema; Type 1 diabetes mellitus with moderate nonproliferative diabetic retinopathy with macular edema, bilateral; Type 1 diabetes mellitus with moderate nonproliferative diabetic retinopathy with macular edema, left eye; Type 1 diabetes mellitus with moderate nonproliferative diabetic retinopathy with macular edema, right eye; Type 1 diabetes mellitus with moderate nonproliferative diabetic retinopathy with macular edema, unspecified eye; Type 1 diabetes mellitus with moderate nonproliferative diabetic retinopathy without macular edema; Type 1 diabetes mellitus with moderate nonproliferative diabetic retinopathy without macular edema, bilateral; Type 1 diabetes mellitus with moderate nonproliferative diabetic retinopathy without macular edema, left eye; Type 1 diabetes mellitus with moderate nonproliferative diabetic retinopathy without macular edema, right eye; Type 1 diabetes mellitus with moderate nonproliferative diabetic retinopathy without macular edema, unspecified eye; Type 1 diabetes mellitus with neurological complications; Type 1 diabetes mellitus with ophthalmic complications; Type 1 diabetes mellitus with oral complications; Type 1 diabetes mellitus with other circulatory complications; Type 1 diabetes mellitus with other diabetic arthropathy; Type 1 diabetes mellitus with other diabetic kidney complication; Type 1 diabetes mellitus with other diabetic neurological complication; Type 1 diabetes mellitus with other diabetic ophthalmic complication; Type 1 diabetes mellitus with other oral complications; Type 1 diabetes mellitus with other skin complications; Type 1 diabetes mellitus with other skin ulcer; Type 1 diabetes mellitus with other specified complication; Type 1 diabetes mellitus with other specified complications; Type 1 diabetes mellitus with proliferative diabetic retinopathy; Type 1 diabetes mellitus with proliferative diabetic retinopathy with combined traction retinal detachment and rhegmatogenous retinal detachment; Type 1 diabetes mellitus with proliferative diabetic retinopathy with combined traction retinal detachment and rhegmatogenous retinal detachment, bilateral; Type 1 diabetes mellitus with proliferative diabetic retinopathy with combined traction retinal detachment and rhegmatogenous retinal detachment, left eye; Type 1 diabetes mellitus with proliferative diabetic retinopathy with combined traction retinal detachment and rhegmatogenous retinal detachment, right eye; Type 1 diabetes mellitus with proliferative diabetic retinopathy with macular edema; Type 1 diabetes mellitus with proliferative diabetic retinopathy with macular edema, bilateral; Type 1 diabetes mellitus with proliferative diabetic retinopathy with macular edema, left eye; Type 1 diabetes mellitus with proliferative diabetic retinopathy with macular edema, right eye; Type 1 diabetes mellitus with proliferative diabetic retinopathy with macular edema, unspecified eye; Type 1 diabetes mellitus with proliferative diabetic retinopathy with traction retinal detachment involving the macula; Type 1 diabetes mellitus with proliferative diabetic retinopathy with traction retinal detachment involving the macula, bilateral; Type 1 diabetes mellitus with proliferative diabetic retinopathy with traction retinal detachment involving the macula, left eye; Type 1 diabetes mellitus with proliferative diabetic retinopathy with traction retinal detachment involving the macula, right eye; Type 1 diabetes mellitus with proliferative diabetic retinopathy with traction retinal detachment not involving the macula; Type 1 diabetes mellitus with proliferative diabetic retinopathy with traction retinal detachment not involving the macula, bilateral; Type 1 diabetes mellitus with proliferative diabetic retinopathy with traction retinal detachment not involving the macula, left eye; Type 1 diabetes mellitus with proliferative diabetic retinopathy with traction retinal detachment not involving the macula, right eye; Type 1 diabetes mellitus with proliferative diabetic retinopathy without macular edema; Type 1 diabetes mellitus with proliferative diabetic retinopathy without macular edema, bilateral; Type 1 diabetes mellitus with proliferative diabetic retinopathy without macular edema, left eye; Type 1 diabetes mellitus with proliferative diabetic retinopathy without macular edema, right eye; Type 1 diabetes mellitus with proliferative diabetic retinopathy without macular edema, unspecified eye; Type 1 diabetes mellitus with severe nonproliferative diabetic retinopathy; Type 1 diabetes mellitus with severe nonproliferative diabetic retinopathy with macular edema; Type 1 diabetes mellitus with severe nonproliferative diabetic retinopathy with macular edema, bilateral; Type 1 diabetes mellitus with severe nonproliferative diabetic retinopathy with macular edema, left eye; Type 1 diabetes mellitus with severe nonproliferative diabetic retinopathy with macular edema, right eye; Type 1 diabetes mellitus with severe nonproliferative diabetic retinopathy with macular edema, unspecified eye; Type 1 diabetes mellitus with severe nonproliferative diabetic retinopathy without macular edema; Type 1 diabetes mellitus with severe nonproliferative diabetic retinopathy without macular edema, bilateral; Type 1 diabetes mellitus with severe nonproliferative diabetic retinopathy without macular edema, left eye; Type 1 diabetes mellitus with severe nonproliferative diabetic retinopathy without macular edema, right eye; Type 1 diabetes mellitus with severe nonproliferative diabetic retinopathy without macular edema, unspecified eye; Type 1 diabetes mellitus with skin complications; Type 1 diabetes mellitus with stable proliferative diabetic retinopathy; Type 1 diabetes mellitus with stable proliferative diabetic retinopathy, bilateral; Type 1 diabetes mellitus with stable proliferative diabetic retinopathy, left eye; Type 1 diabetes mellitus with stable proliferative diabetic retinopathy, right eye; Type 1 diabetes mellitus with stable proliferative diabetic retinopathy, unspecified eye; Type 1 diabetes mellitus with unspecified complications; Type 1 diabetes mellitus with unspecified diabetic retinopathy; Type 1 diabetes mellitus with unspecified diabetic retinopathy with macular edema; Type 1 diabetes mellitus with unspecified diabetic retinopathy without macular edema; Type 1 diabetes mellitus without complications

**hypertension** Source Concepts: Benign essential hypertension; Essential (primary) hypertension; Essential hypertension; Unspecified essential hypertension

**height** Body height

**bmi** field ID: Body mass index; Body mass index (BMI) [Ratio]

**total bilirubin** Bilirubin.total [Mass/volume] in Serum or Plasma

**hdl** Cholesterol in HDL [Mass/volume] in Serum or Plasma

### 2 Polygenic score computation

Here we provide additional details, including mathematical descriptions, of how to generate the polygenic scores described in the main text.

In step (2) we build residual phenotypes. For a raw phenotype, $\vec{y}$, We regress

$$\vec{y} \sim \vec{\theta} \cdot \bar{K} \to \vec{\theta}^* \,, \tag{2.1}$$

for covariate matrix $\bar{K}$ and coefficients $\vec{\theta}$. Residual phenotypes, $\vec{y}'$ are then built by subtracting off the covariate contribution from the raw (or sex specific z-scored) phenotype

$$\vec{y}' = \vec{y} - \vec{y}^* \qquad \text{where} \qquad \vec{y}^* = \vec{\theta}^* \cdot \bar{K} \,. \tag{2.2}$$

In step (4) we use Scikit-Learn to train predictors. This is a straightforward application of using coordinate descent to minimize the LASSO objective function,

$$\mathcal{O}(\lambda) = \frac{1}{2N} ||\vec{y}' - \bar{X} \cdot \vec{\beta}||^2_{L_2} + \lambda |\vec{\beta}|_{L_1} \,, \tag{2.3}$$

for features across the entire autosome or on the blocks (chromosome). Here $\bar{X}$ is the encoded genotype matrix, $N$ is the number of samples (i.e., the length of $\vec{y}$), and $\vec{\beta}$ are the SNV weights. As detailed in the associated code examples, we used the Scikit-Learn function `linear_model.lasso_path` with 100-200 steps, $0.0005 < \lambda_{min}/\lambda_{max} < 0.001$, and let it run for a maximum of 1500 iterations. Examples of the validation paths for block vs global LASSO in the UKB can be found in the section 6. Model selection, $\lambda \to \lambda^*$, is done by selecting the maximal performance in the validation/model-selection set.

In step (5) we perform the block regression. We index each block by the label $b$. For each block we have a genotype matrix, $X^b_{ij}$, where the $i$ indexes the samples and $j$ the features. Unweighted scores for each block can be written as: $\sum_j \bar{X}^b_{ij} \cdot \beta^b_j$. For a phenotype, $y_i$, we determine the individual block weights, $\alpha_b$, via linear regression

$$y_j \sim \sum_b \alpha_b \times \sum_i \bar{X}^b_{ij} \cdot \beta^b_j \to \alpha^* \,. \tag{2.4}$$

The final step (6) involves constructing the full model by applying

$$\mathcal{B}^b_j = \alpha_b \cdot \beta^b_j \tag{2.5}$$

to genotypes.

#### 2.1 Uncertainty

Error bars in plots include a contribution from finite sample sizes using the following definitions:

$$\sigma_{\text{AUC}} \approx \sqrt{\left(\frac{1}{3\sqrt{N_{cases}}}\right)^2 + \sigma^2_{cv-SD}} \qquad \text{and} \qquad \sigma_{corr.} = \sqrt{\left(\sqrt{\frac{1-\rho^2}{N-2}}\right)^2 + \sigma^2_{cv-SD}}\,, \tag{2.6}$$

where $N_{cases}$ is the number of cases used in training, $\rho$ is the correlation, $N$ is the total number of samples (people) used in training, and $\sigma_{cv-SD}$ is the standard deviation over cross-validation folds. The first factor in the AUC uncertainty is empirically determined in [1].

##### 2.2 PGS variance distributions

We can approximate the variance attributed to each region in the genome as follows. Within each block, the encoded genotype can be written ${}^{b}X_{i,j}$ where $b$ labels the block, $i$ labels the sample, and $j$ labels the feature (SNV). We then include the weights for each feature, ${}^{b}\beta_j$, and each block, $\alpha_b$: $\alpha_b\,{}^{b}\beta_j\,{}^{b}X_{i,j} \equiv {}^{b}H_{i,j}$. Finally we compute the covariance matrix

$$K_{j,k} = COV(\vec{H}_j, \vec{H}_k)\,, \tag{2.7}$$

where $\vec{H}_j$ is the column of sample values for the $j$th feature. Note that when computing the covariance between features (SNVs) on different blocks the covariance is 0 by definition.

##### 2.3 Computational resource management

Computational resources used in both the blockLASSO and global approach were monitored for the UKB calculations. The computing cluster used for this analysis uses a SLURM [2] workload manager. The results shown in the main text were tracked via the `seff` command.

#### 3 PGS metrics plots

More comparisons of block vs global LASSO in AoU and UKB* (UKB reduced to training/testing sets comparable to AoU) can be seen in **Figure 1** - **Figure 3**. Uncertainties reflect one standard deviation computed from 5-fold cross-validation and computing AUC/correlation with finite sample sizes (effects are added in quadrature). For the "global AoU" measurement only one training fold was run so the uncertainty is the larger of the finite size effect, or the corresponding uncertainty found in the UKB.

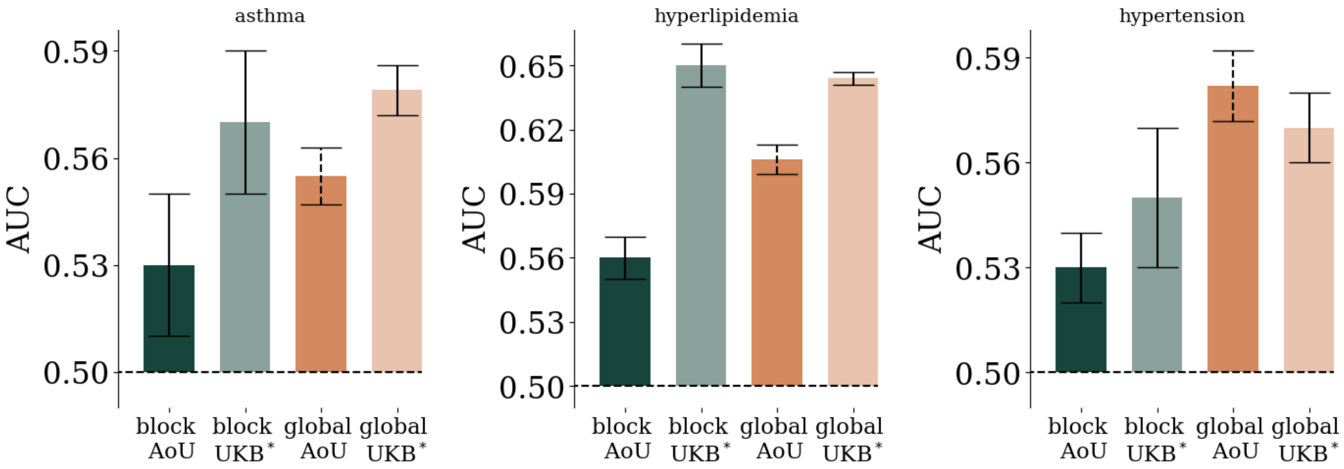

**Figure 1:** Block vs global lasso performance in AoU and UKB* for asthma, hyperlipidemia, and hypertension.

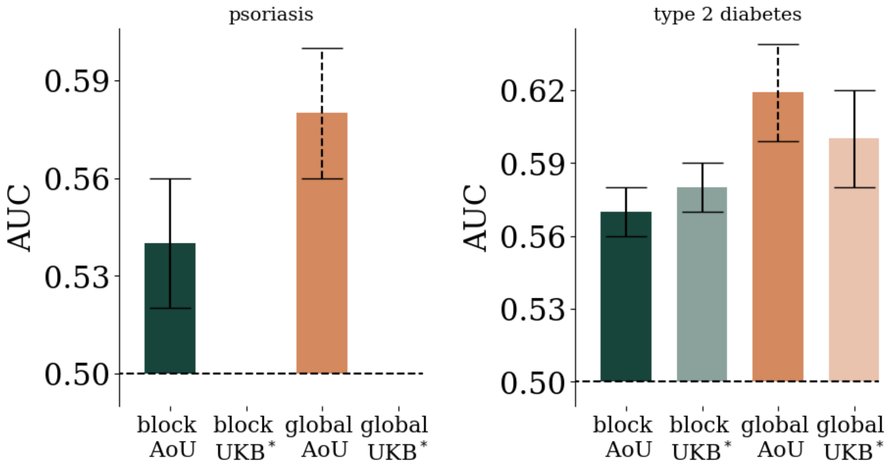

**Figure 2:** Block vs global lasso performance in AoU and UKB* for psoriasis and type 2 diabetes. Because there are more cases in AoU than the UKB, a UKB* set was not trained for psoriasis.

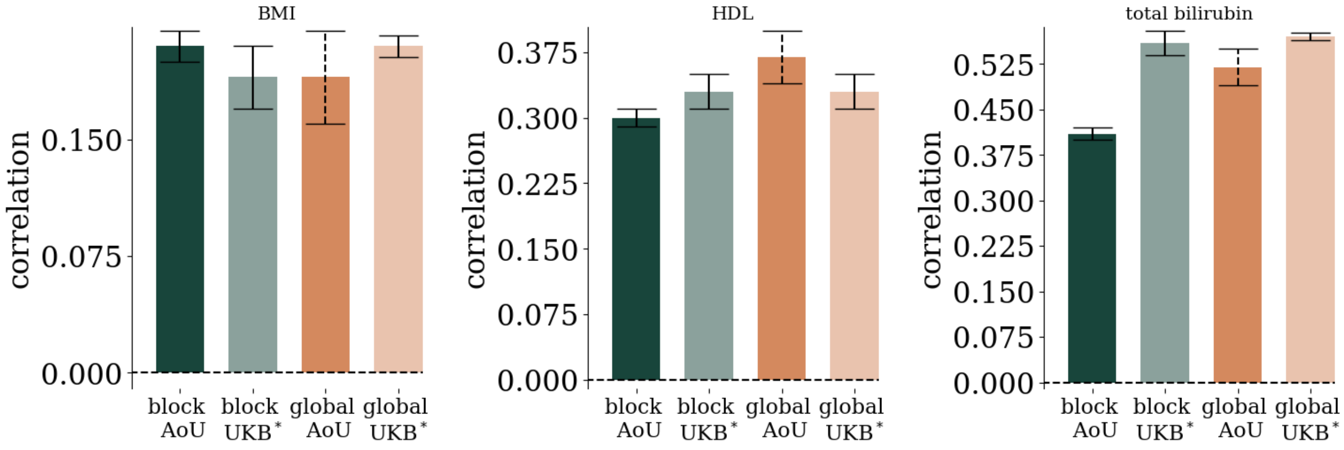

**Figure 3:** Block vs global lasso performance in AoU and UKB* for bmi, hdl, and total bilirubin.

### 4 Variance plots

Here we report the additional variance explained per approximate base position and per chromosome, **Figure 4 - Figure 13**, for the phenotypes not displayed in the main text. The variance per location is normalized by the total variance explained by the PGS. The variance per location is filtered to only include contributions of at least 0.01% of the total PGS varaince. To make the variance per base pair position human readable we use 1 megabase pair sized bins.

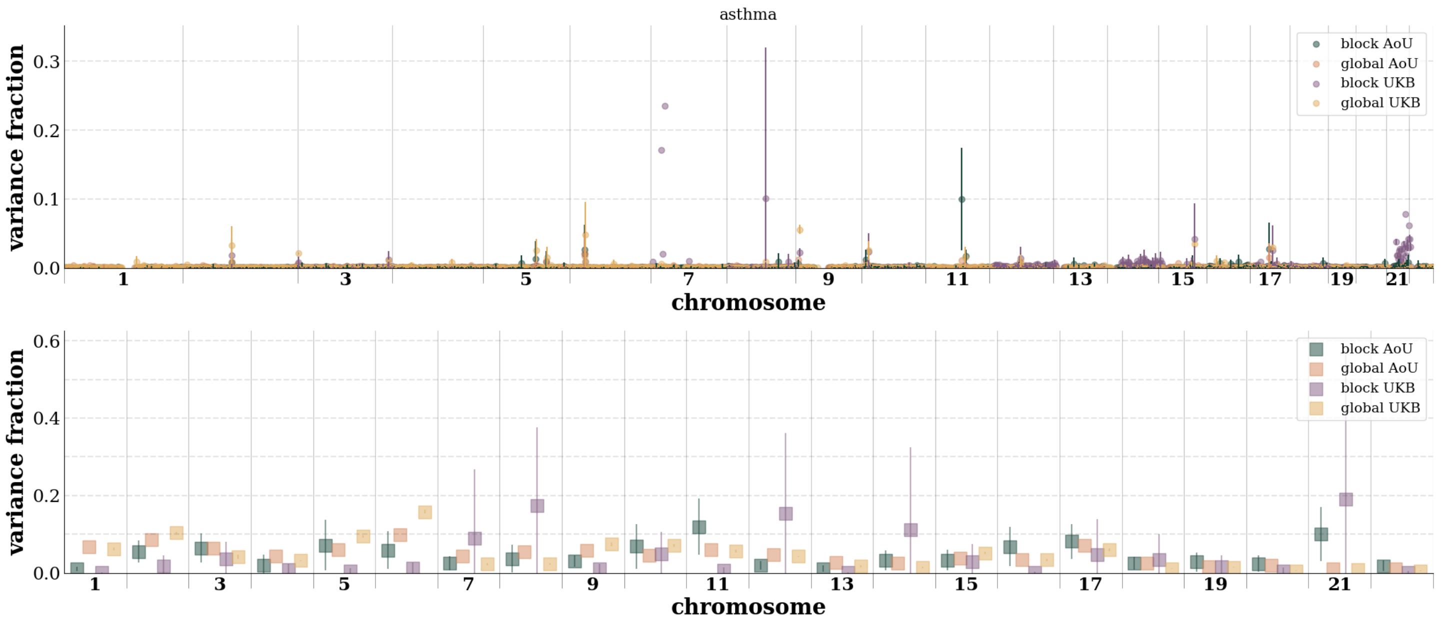

**Figure 4:** Fraction of variance explained per binned base pair position (top) and per chromosome (bottom) in AoU and the UKB for asthma.

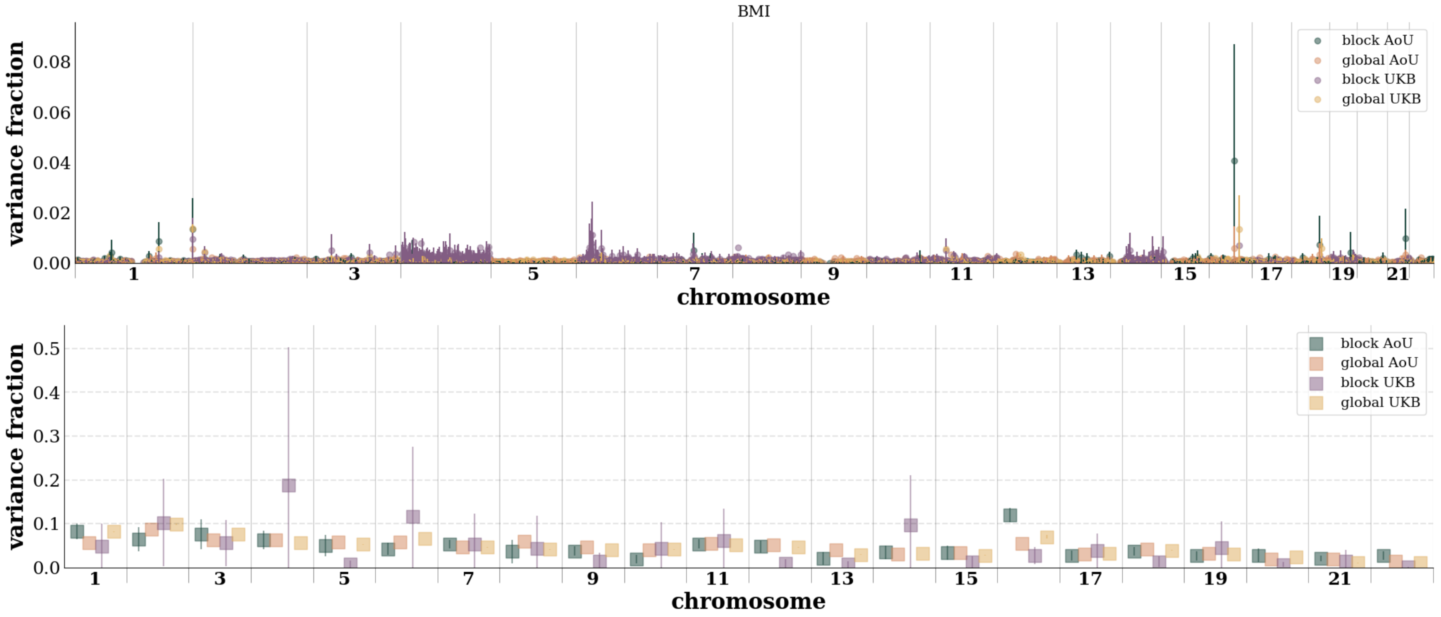

**Figure 5:** Fraction of variance explained per binned base pair position (top) and per chromosome (bottom) in AoU and the UKB for bmi.

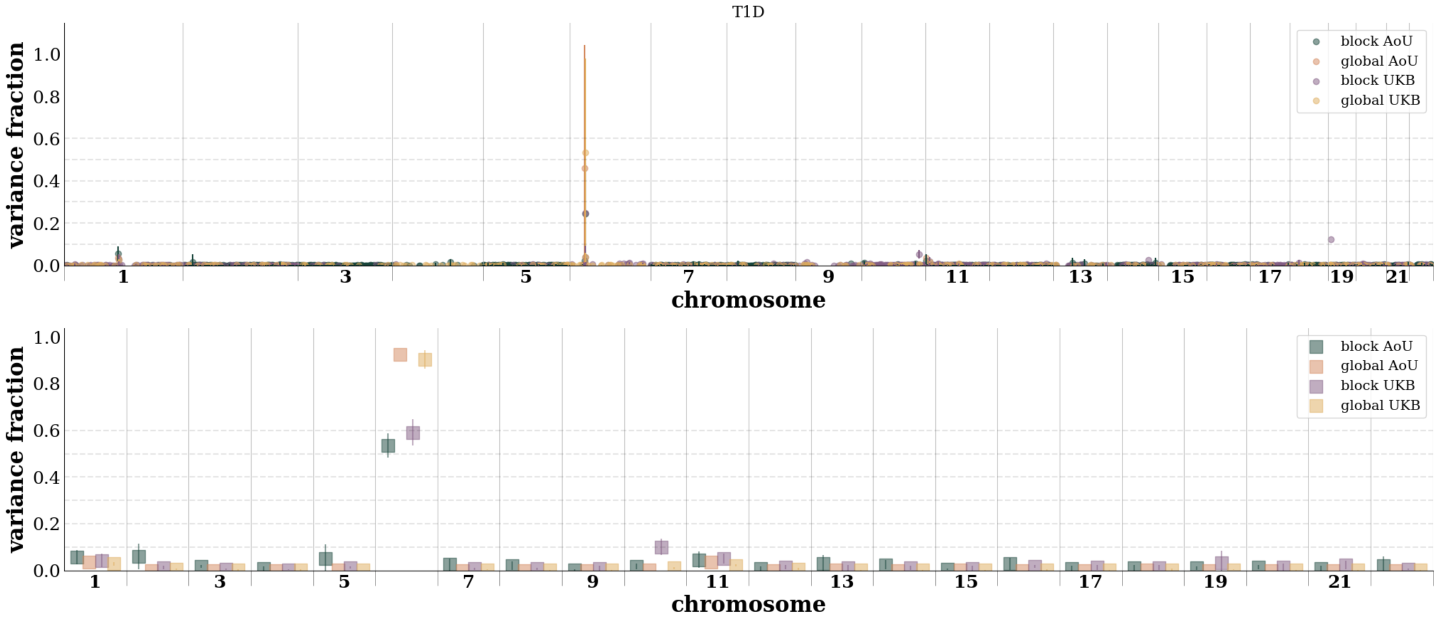

**Figure 6:** Fraction of variance explained per binned base pair position (top) and per chromosome (bottom) in AoU and the UKB for type 1 diabetes.

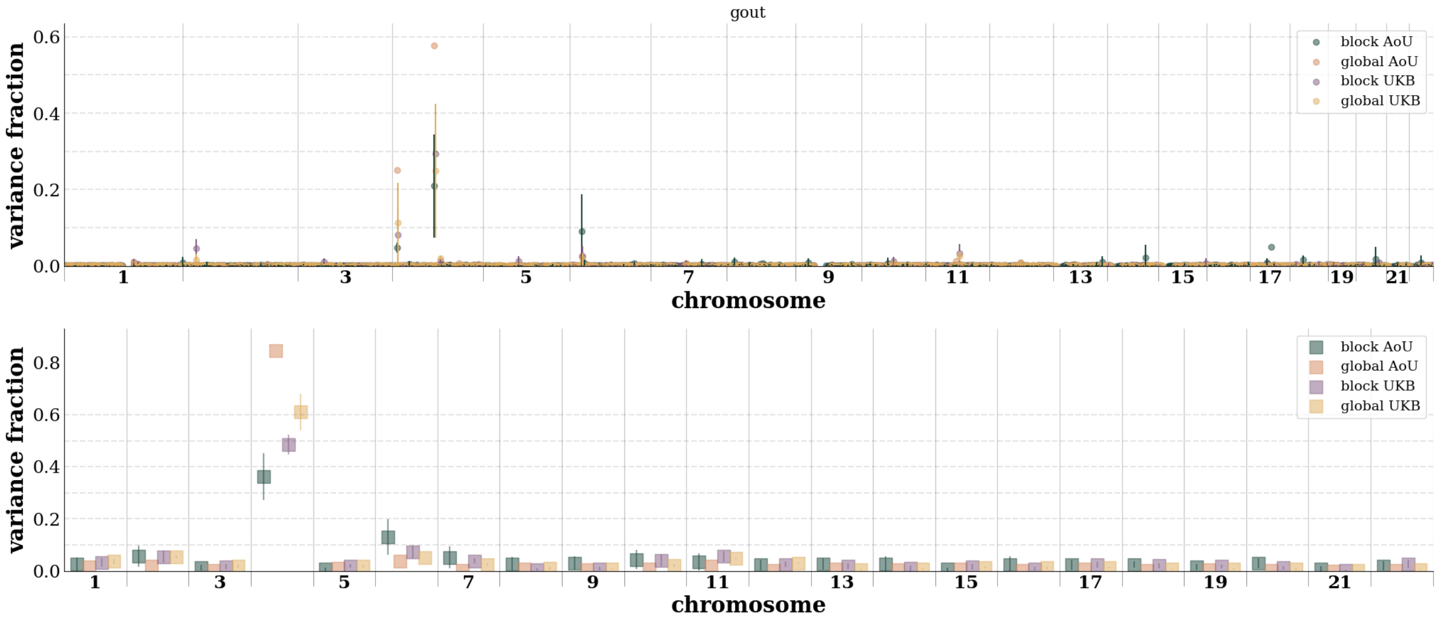

**Figure 7:** Fraction of variance explained per binned base pair position (top) and per chromosome (bottom) in AoU and the UKB for gout.

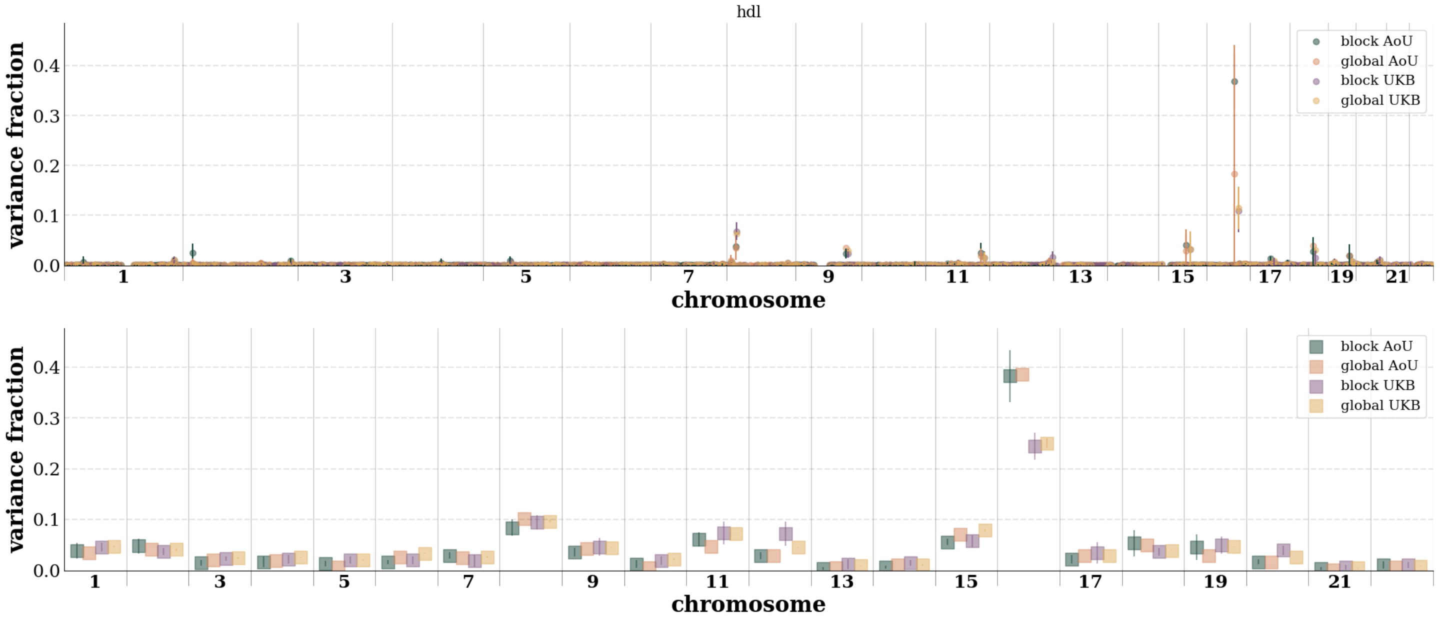

**Figure 8:** Fraction of variance explained per binned base pair position (top) and per chromosome (bottom) in AoU and the UKB for hdl.

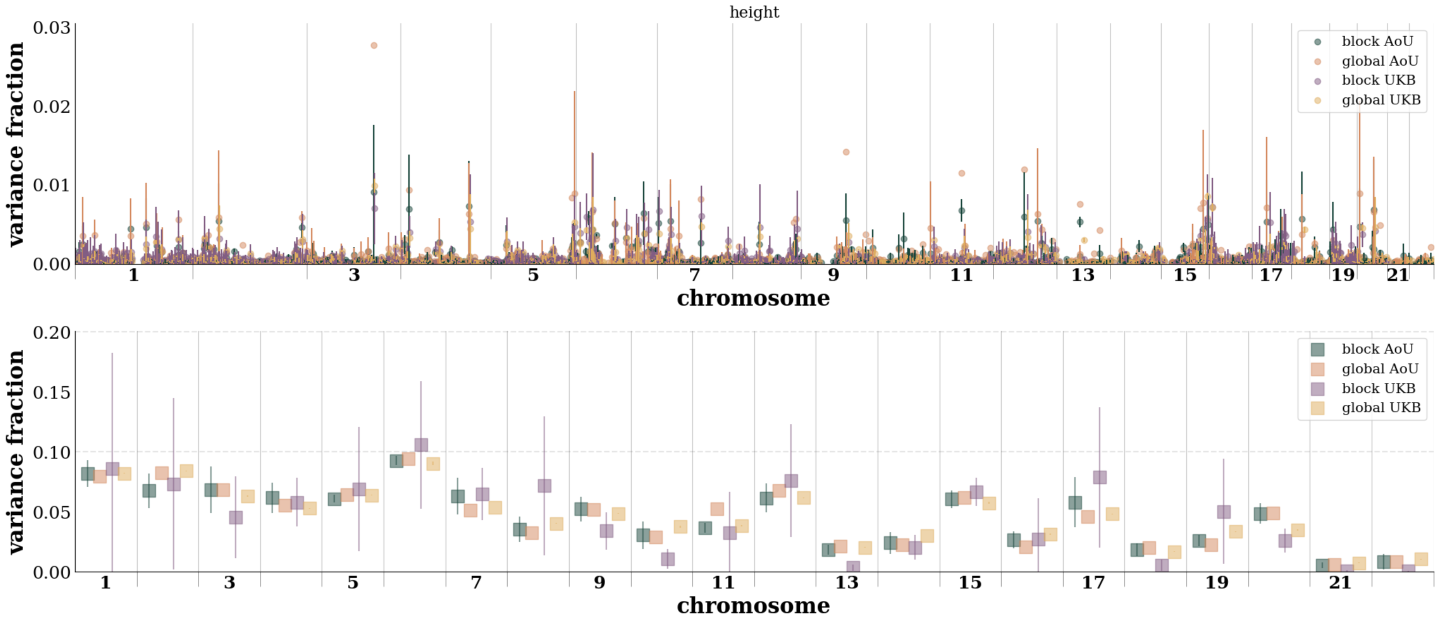

**Figure 9:** Fraction of variance explained per binned base pair position (top) and per chromosome (bottom) in AoU and the UKB for height.

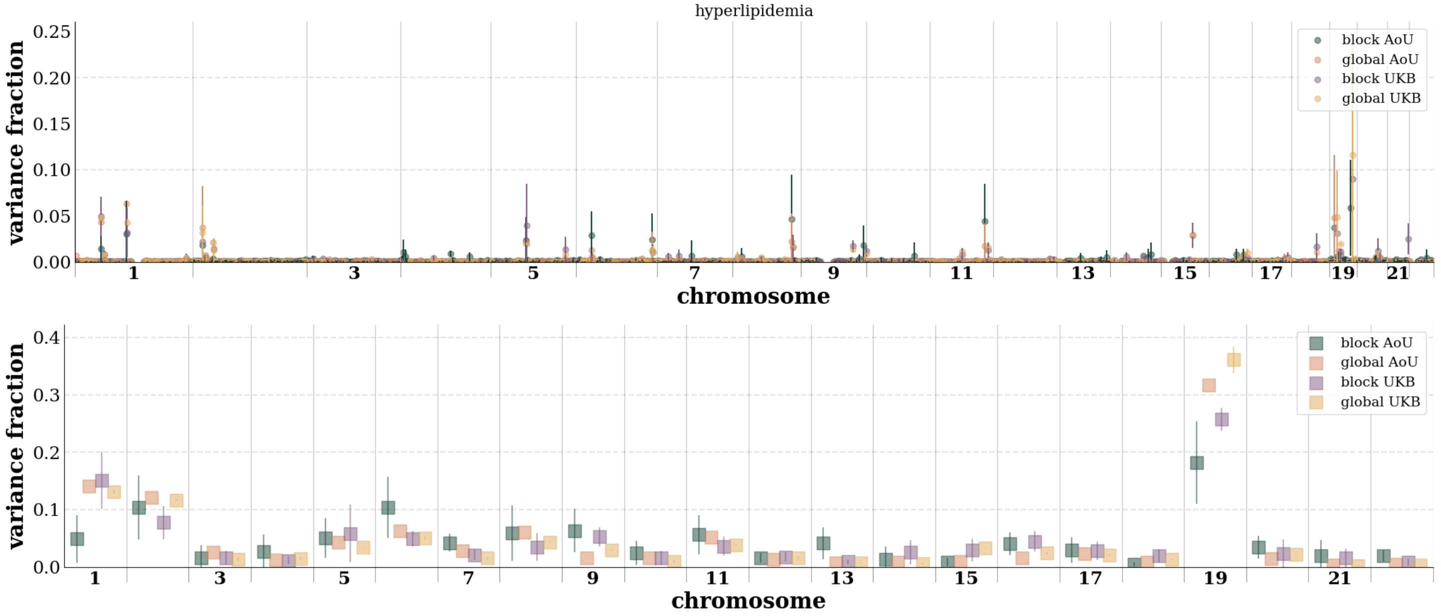

**Figure 10:** Fraction of variance explained per binned base pair position (top) and per chromosome (bottom) in AoU and the UKB for hyperlipidemia.

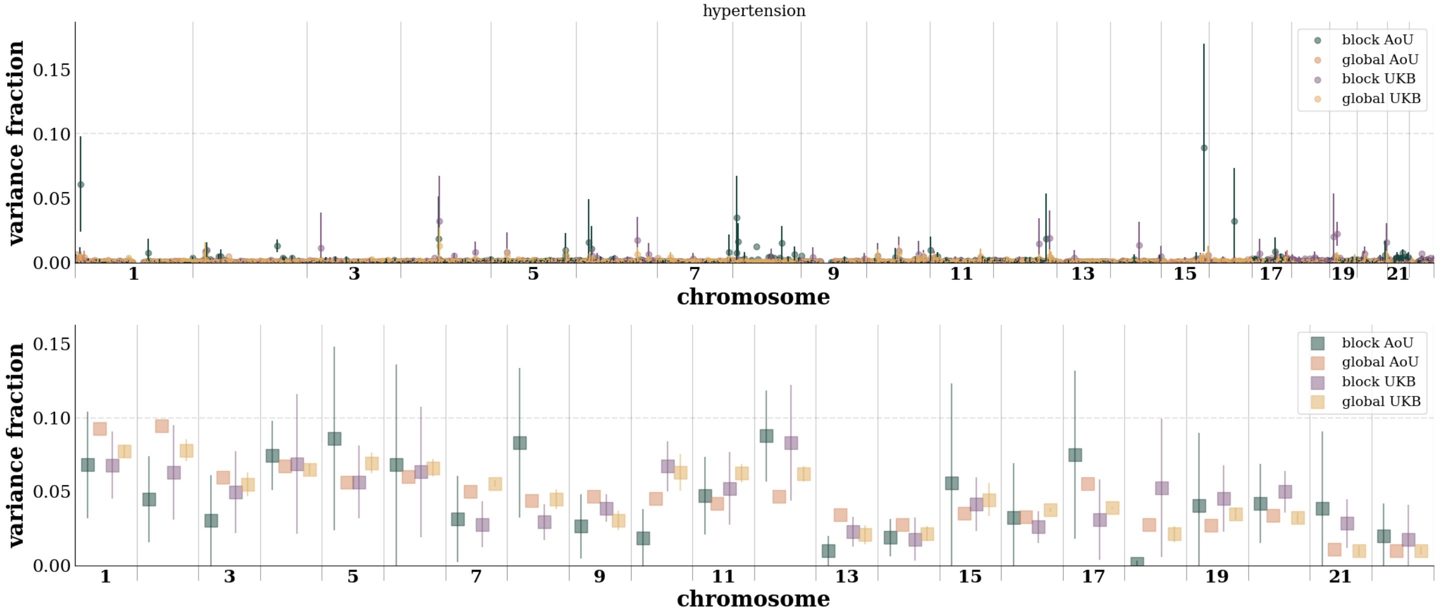

**Figure 11:** Fraction of variance explained per binned base pair position (top) and per chromosome (bottom) in AoU and the UKB for hypertension.

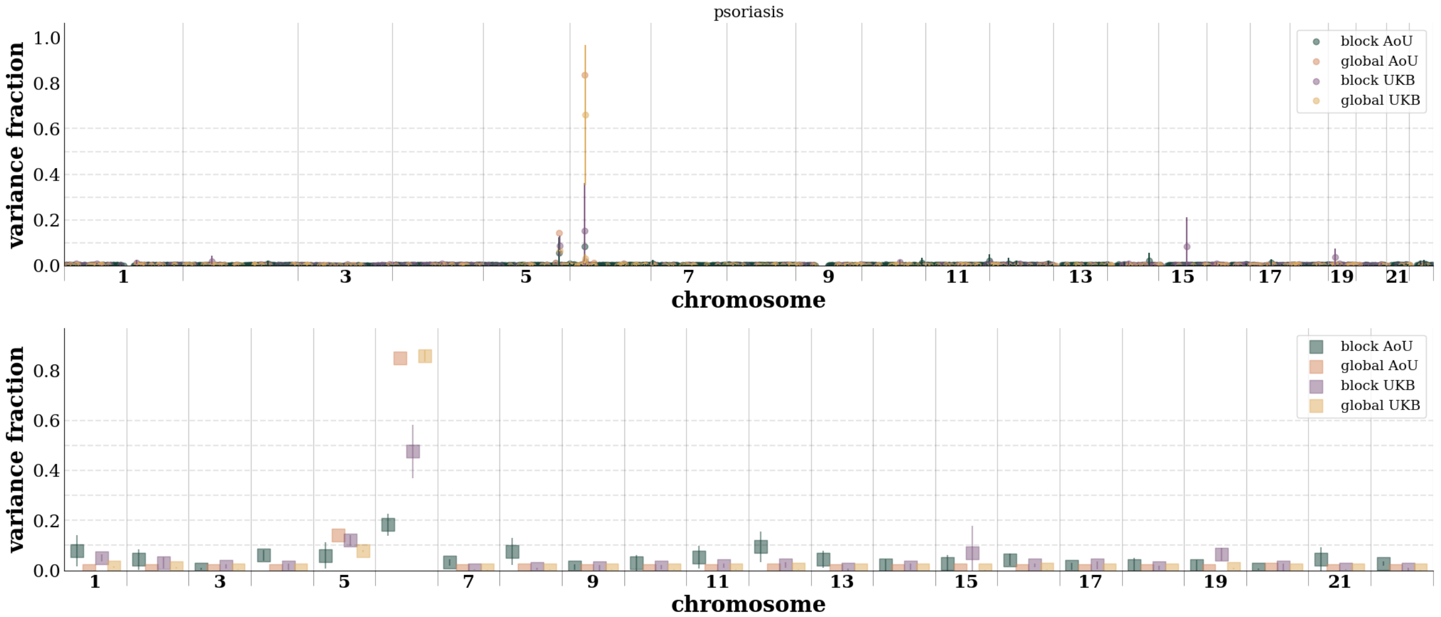

**Figure 12:** Fraction of variance explained per binned base pair position (top) and per chromosome (bottom) in AoU and the UKB for psoriasis.

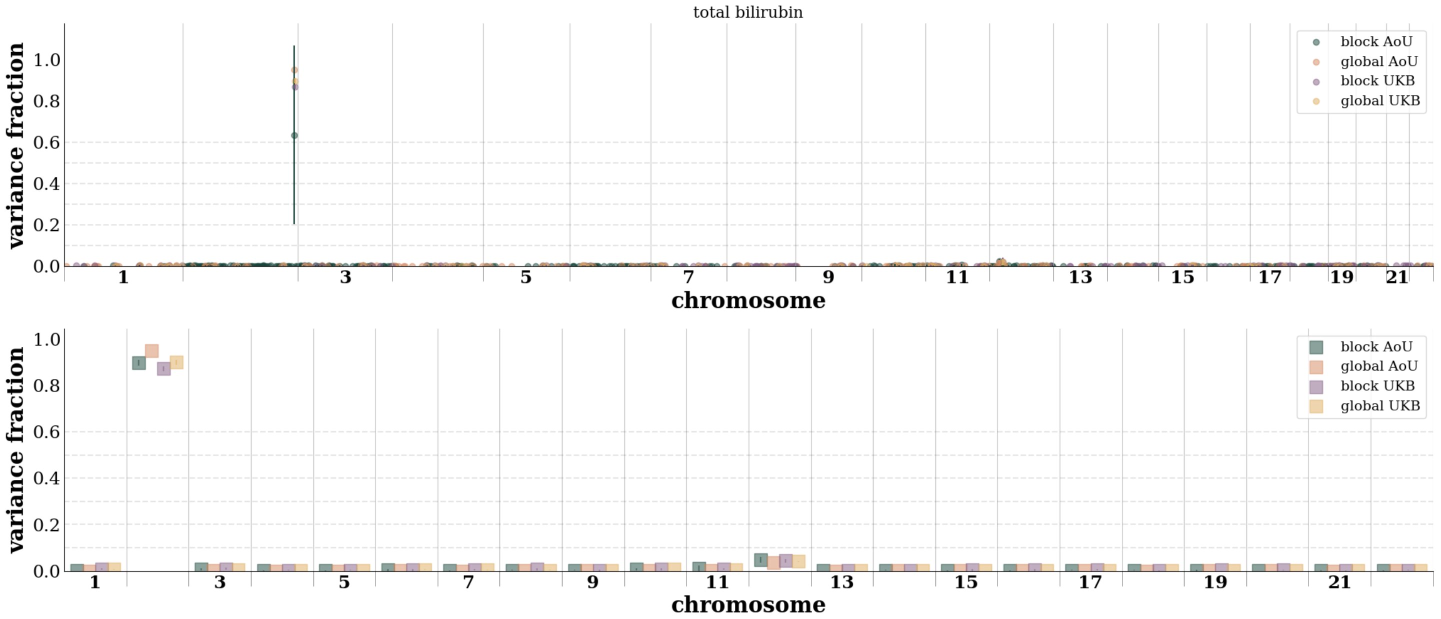

**Figure 13:** Fraction of variance explained per binned base pair position (top) and per chromosome (bottom) in AoU and the UKB for total bilirubin.

### 5 Training and re-weighting

Here we show the additional plots which demonstrate performance as a function of training block sizes and the effect of the re-weighting the blocks: **Figure 14 - Figure 23**. Different training sizes – {10; 23; 50; 100; 227; 500; 1,000; 2,273; 5,000; 10,000; 22,727} SNVs – were tested in the UKB. The results for the different block sizes included the block re-weighting. All training and re-weighting was done with the EUR populations. Examples of the effects of re-weighting on all populations used from AoU can be seen in this section.

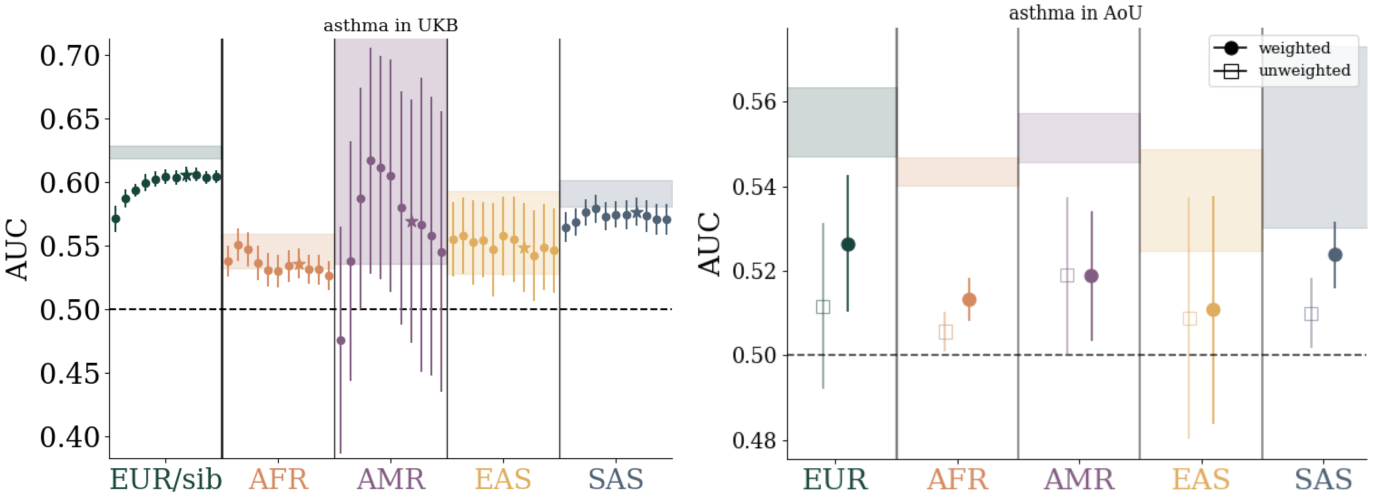

**Figure 14:** Left: performance as a function of training SNV size in UKB and applied to different ancestry groups. Within each ancestry group, dots correspond to training with $\{10, 23, 50, 100, 227, 500, 1000, 2273, 5000, 10000, 22727\}$ SNVs per chromosome from left to right respectively. The starred data points correspond to 2273 SNVs per chromosome wich is roughly equivalent to 50k SNVs across the autosome. Right: performance before and after the re-weighting step of the blockLASSO. While re-weighting is trained within the EUR group, the effect of re-weighting improves prediction accuracy across all tested ancestry groupings.

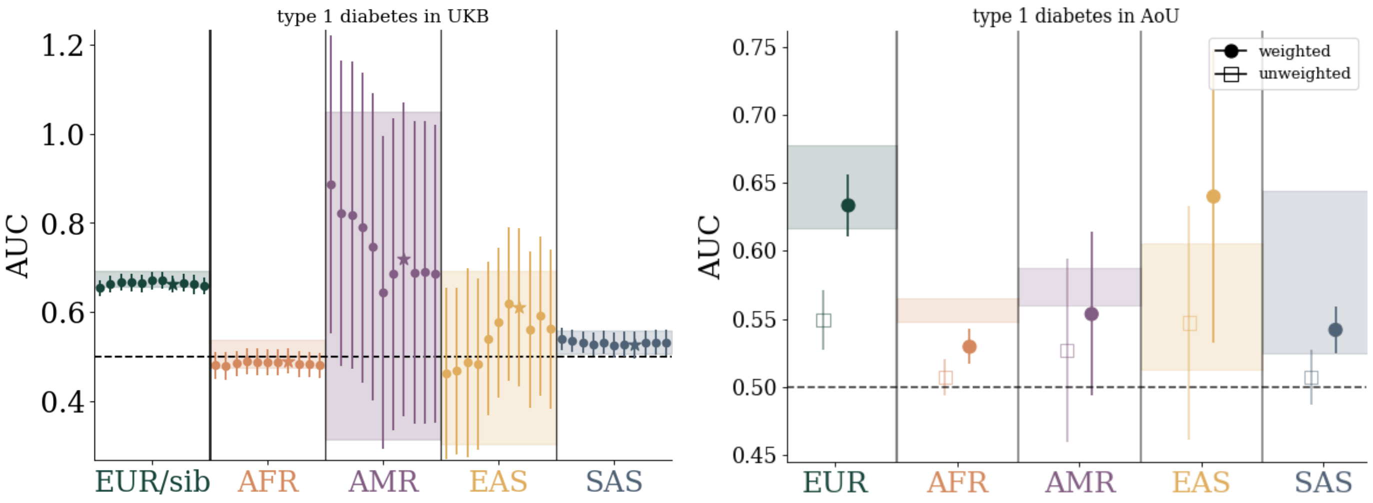

**Figure 15:** Left: performance as a function of training SNV size in UKB and applied to different ancestry groups. Within each ancestry group, dots correspond to training with $\{10, 23, 50, 100, 227, 500, 1000, 2273, 5000, 10000, 22727\}$ SNVs per chromosome from left to right respectively. The starred data points correspond to 2273 SNVs per chromosome wich is roughly equivalent to 50k SNVs across the autosome. Right: performance before and after the re-weighting step of the blockLASSO. While re-weighting is trained within the EUR group, the effect of re-weighting improves prediction accuracy across all tested ancestry groupings.

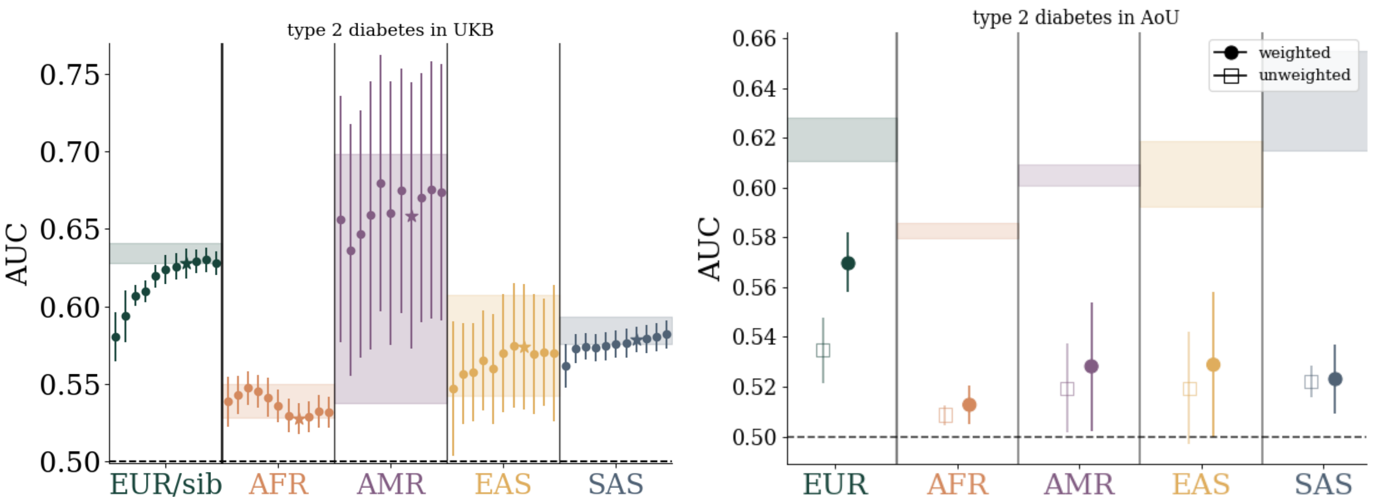

**Figure 16:** Left: performance as a function of training SNV size in UKB and applied to different ancestry groups. Within each ancestry group, dots correspond to training with $\{10, 23, 50, 100, 227, 500, 1000, 2273, 5000, 10000, 22727\}$ SNVs per chromosome from left to right respectively. The starred data points correspond to 2273 SNVs per chromosome wich is roughly equivalent to 50k SNVs across the autosome. Right: performance before and after the re-weighting step of the blockLASSO. While re-weighting is trained within the EUR group, the effect of re-weighting improves prediction accuracy across all tested ancestry groupings.

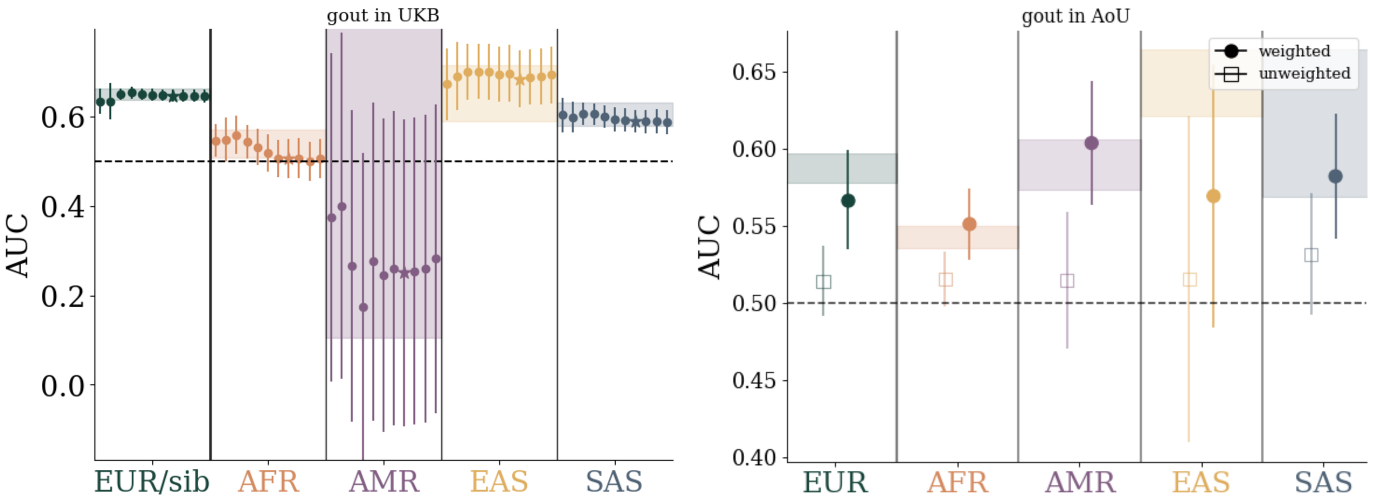

**Figure 17:** Left: performance as a function of training SNV size in UKB and applied to different ancestry groups. Within each ancestry group, dots correspond to training with $\{10, 23, 50, 100, 227, 500, 1000, 2273, 5000, 10000, 22727\}$ SNVs per chromosome from left to right respectively. The starred data points correspond to 2273 SNVs per chromosome wich is roughly equivalent to 50k SNVs across the autosome. Right: performance before and after the re-weighting step of the blockLASSO. While re-weighting is trained within the EUR group, the effect of re-weighting improves prediction accuracy across all tested ancestry groupings.

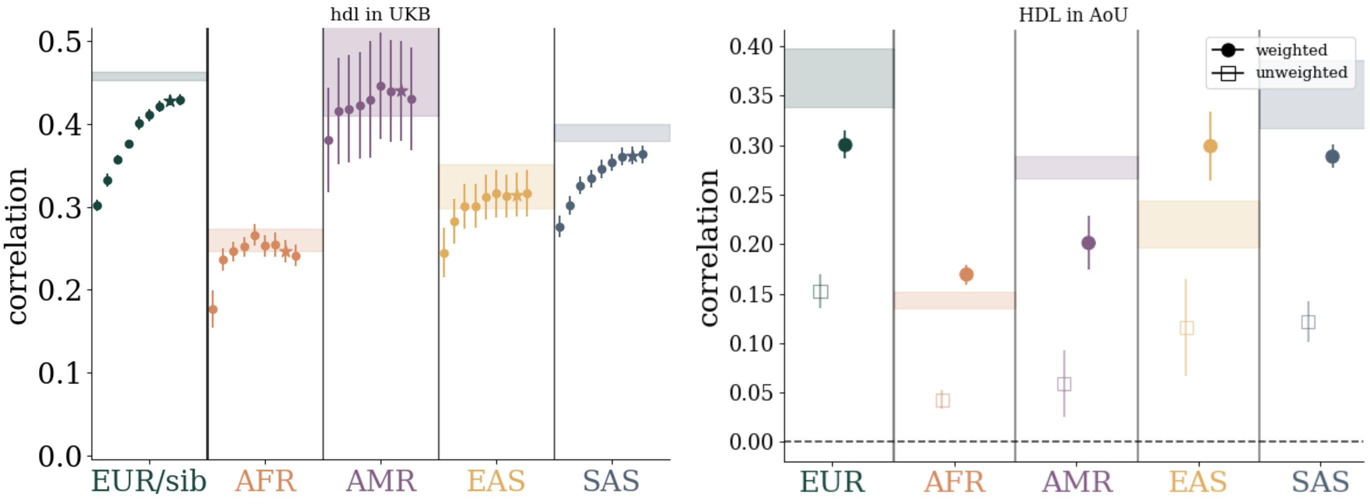

**Figure 18:** Left: performance as a function of training SNV size in UKB and applied to different ancestry groups. Within each ancestry group, dots correspond to training with $\{10, 23, 50, 100, 227, 500, 1000, 2273, 5000, 10000, 22727\}$ SNVs per chromosome from left to right respectively. The starred data points correspond to 2273 SNVs per chromosome wich is roughly equivalent to 50k SNVs across the autosome. Right: performance before and after the re-weighting step of the blockLASSO. While re-weighting is trained within the EUR group, the effect of re-weighting improves prediction accuracy across all tested ancestry groupings.

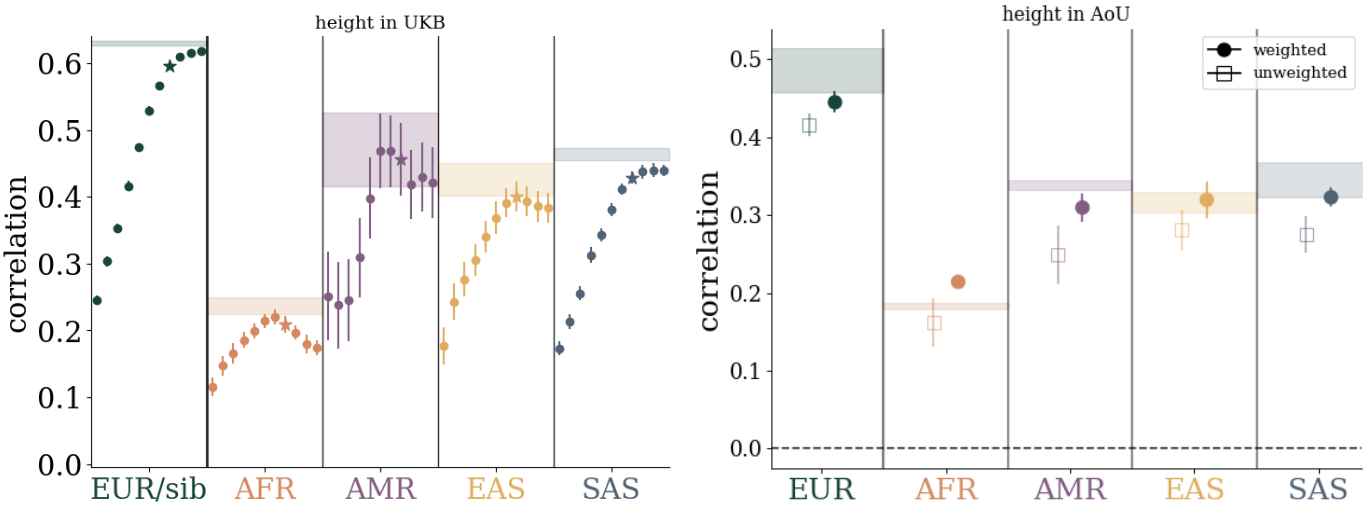

**Figure 19:** Left: performance as a function of training SNV size in UKB and applied to different ancestry groups. Within each ancestry group, dots correspond to training with $\{10, 23, 50, 100, 227, 500, 1000, 2273, 5000, 10000, 22727\}$ SNVs per chromosome from left to right respectively. The starred data points correspond to 2273 SNVs per chromosome wich is roughly equivalent to 50k SNVs across the autosome. Right: performance before and after the re-weighting step of the blockLASSO. While re-weighting is trained within the EUR group, the effect of re-weighting improves prediction accuracy across all tested ancestry groupings.

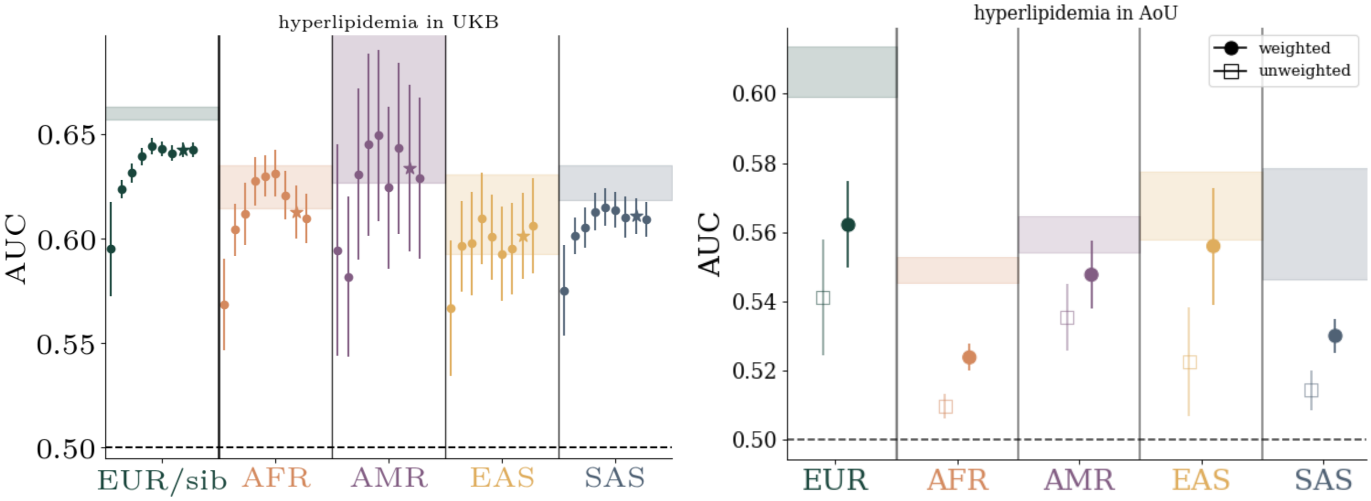

**Figure 20:** Left: performance as a function of training SNV size in UKB and applied to different ancestry groups. Within each ancestry group, dots correspond to training with $\{10, 23, 50, 100, 227, 500, 1000, 2273, 5000, 10000, 22727\}$ SNVs per chromosome from left to right respectively. The starred data points correspond to 2273 SNVs per chromosome wich is roughly equivalent to 50k SNVs across the autosome. Right: performance before and after the re-weighting step of the blockLASSO. While re-weighting is trained within the EUR group, the effect of re-weighting improves prediction accuracy across all tested ancestry groupings.

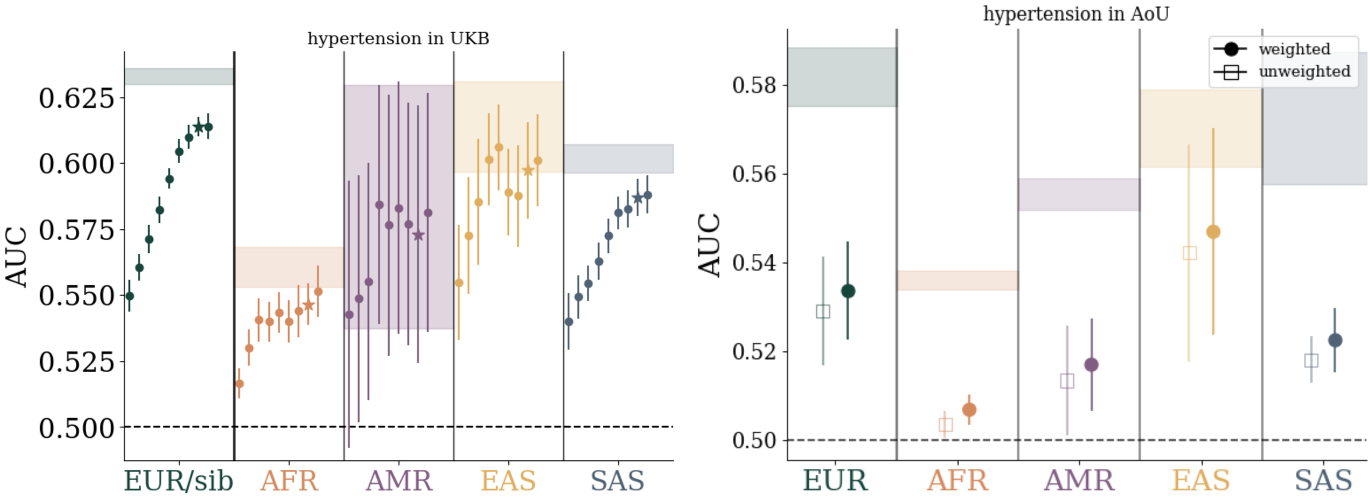

**Figure 21:** Left: performance as a function of training SNV size in UKB and applied to different ancestry groups. Within each ancestry group, dots correspond to training with $\{10, 23, 50, 100, 227, 500, 1000, 2273, 5000, 10000, 22727\}$ SNVs per chromosome from left to right respectively. The starred data points correspond to 2273 SNVs per chromosome wich is roughly equivalent to 50k SNVs across the autosome. Right: performance before and after the re-weighting step of the blockLASSO. While re-weighting is trained within the EUR group, the effect of re-weighting improves prediction accuracy across all tested ancestry groupings.

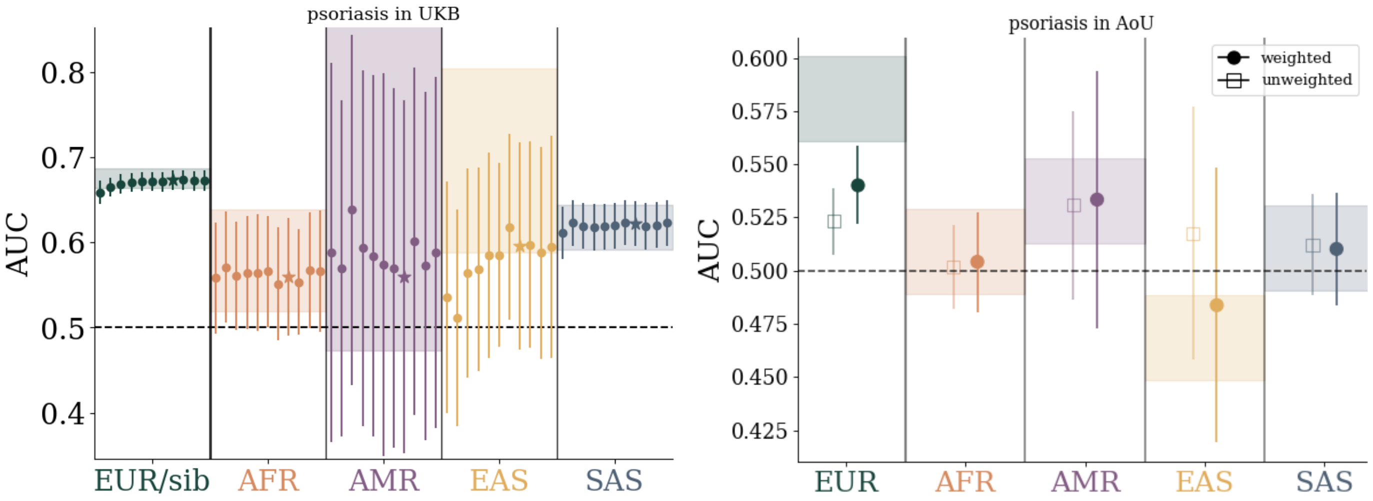

**Figure 22:** Left: performance as a function of training SNV size in UKB and applied to different ancestry groups. Within each ancestry group, dots correspond to training with $\{10, 23, 50, 100, 227, 500, 1000, 2273, 5000, 10000, 22727\}$ SNVs per chromosome from left to right respectively. The starred data points correspond to 2273 SNVs per chromosome wich is roughly equivalent to 50k SNVs across the autosome. Right: performance before and after the re-weighting step of the blockLASSO. While re-weighting is trained within the EUR group, the effect of re-weighting improves prediction accuracy across all tested ancestry groupings.

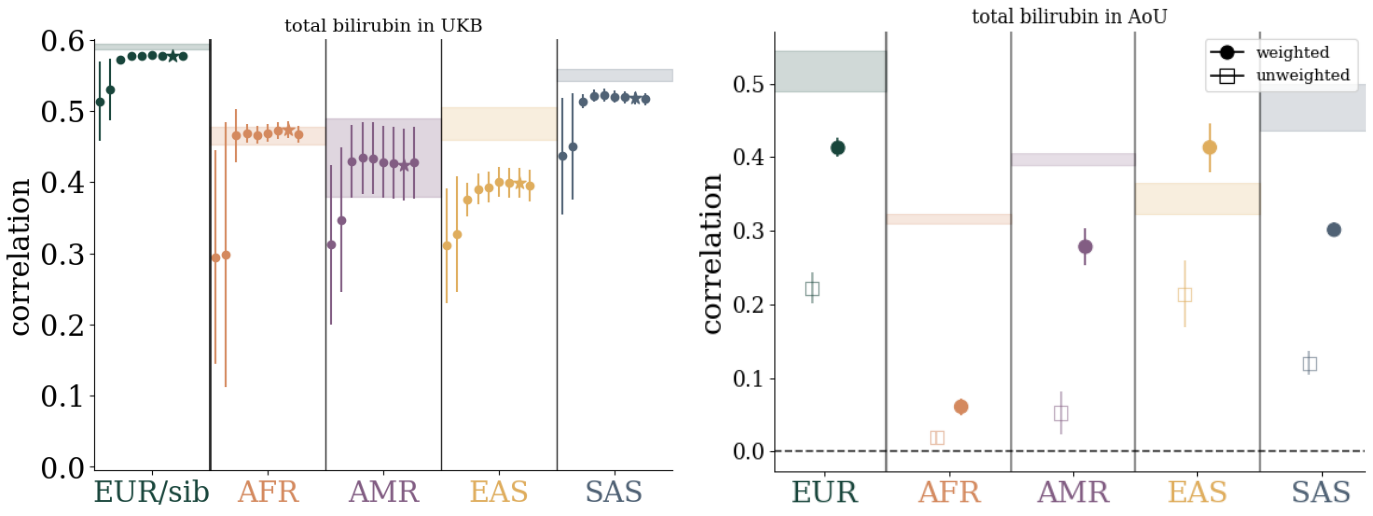

**Figure 23:** Left: performance as a function of training SNV size in UKB and applied to different ancestry groups. Within each ancestry group, dots correspond to training with $\{10, 23, 50, 100, 227, 500, 1000, 2273, 5000, 10000, 22727\}$ SNVs per chromosome from left to right respectively. The starred data points correspond to 2273 SNVs per chromosome wich is roughly equivalent to 50k SNVs across the autosome. Right: performance before and after the re-weighting step of the blockLASSO. While re-weighting is trained within the EUR group, the effect of re-weighting improves prediction accuracy across all tested ancestry groupings.

### 6 LASSO validation paths

Here we show, in **Figure 24 - Figure 34**, the LASSO training paths for the blockLASSO applied to the validation/model-selection sets. The right most column compares the combined and re-weighted blockLASSO result, shaded band, to the global LASSO path.

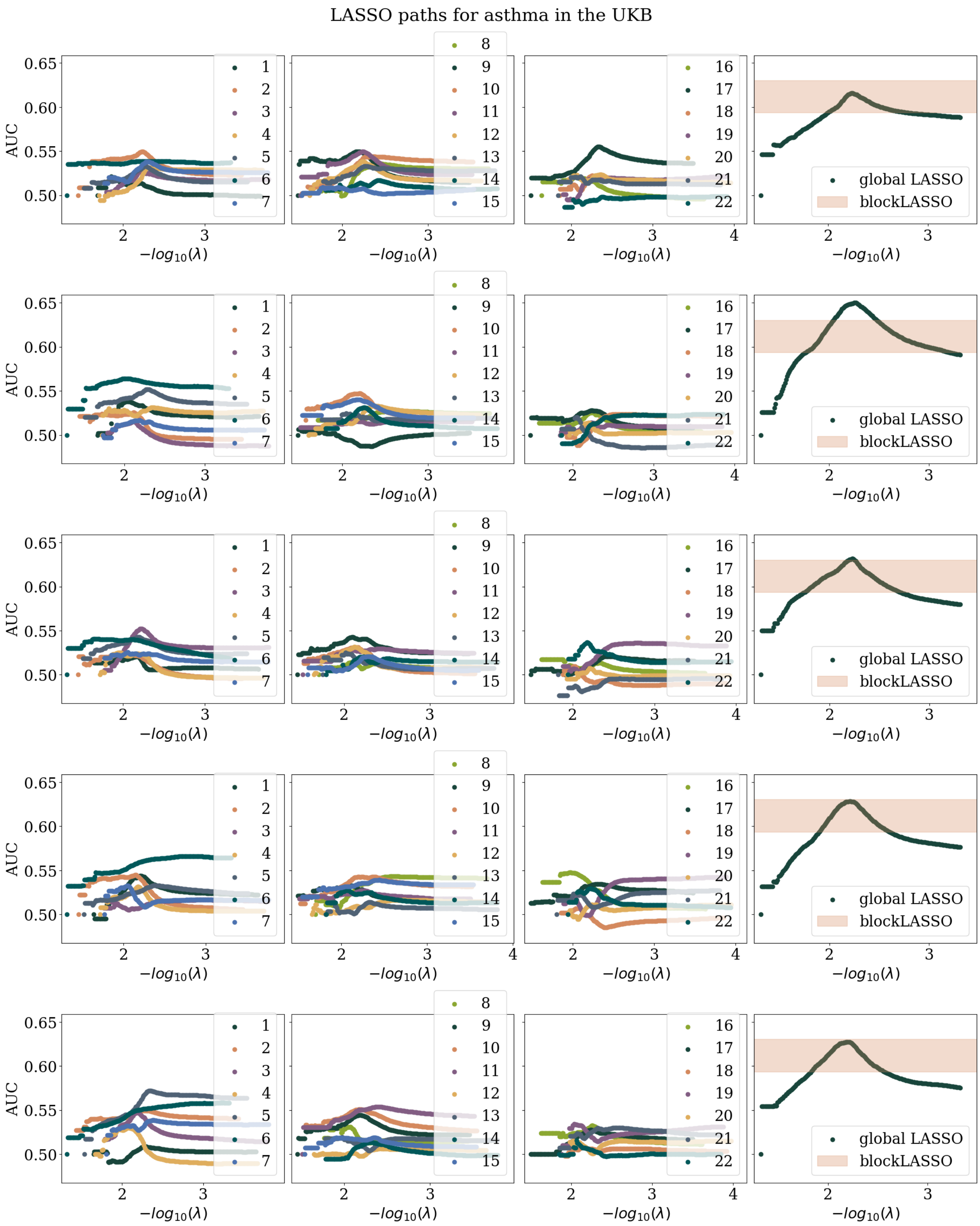

**Figure 24:** Coordinate descent based LASSO paths within the validation/model-selction set within the UKB. The first three panels left to right show the paths for each chromosome for the blockLASSO construction. The fourth panel on the right shows the comparison of the global LASSO (dots) vs the complete (i.e., re-weighted) blockLASSO value in the validation set. Different rows correspond to different cross-validation folds.

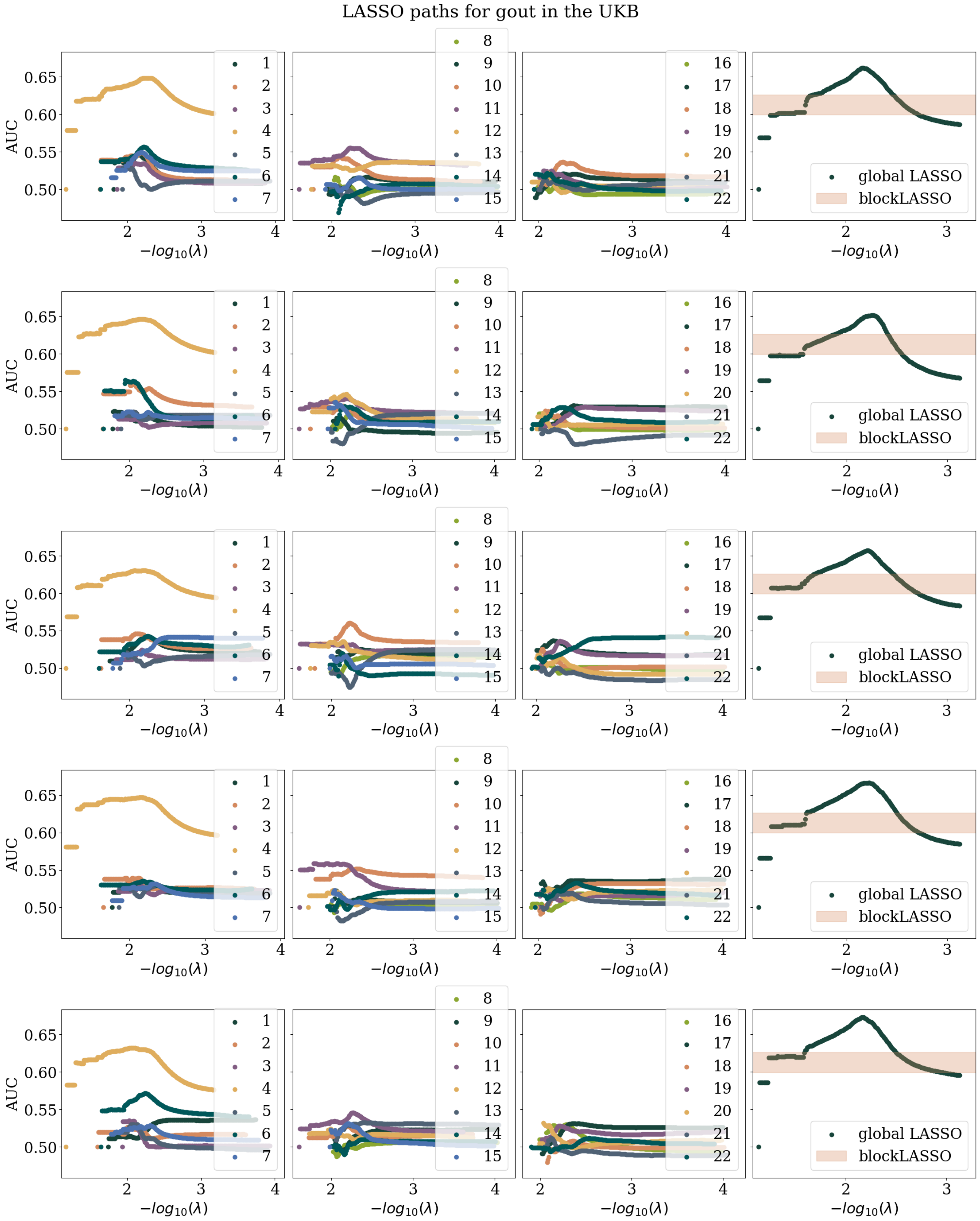

**Figure 25:** Coordinate descent based LASSO paths within the validation/model-selction set within the UKB. The first three panels left to right show the paths for each chromosome for the blockLASSO construction. The fourth panel on the right shows the comparison of the global LASSO (dots) vs the complete (i.e., re-weighted) blockLASSO value in the validation set. Different rows correspond to different cross-validation folds.

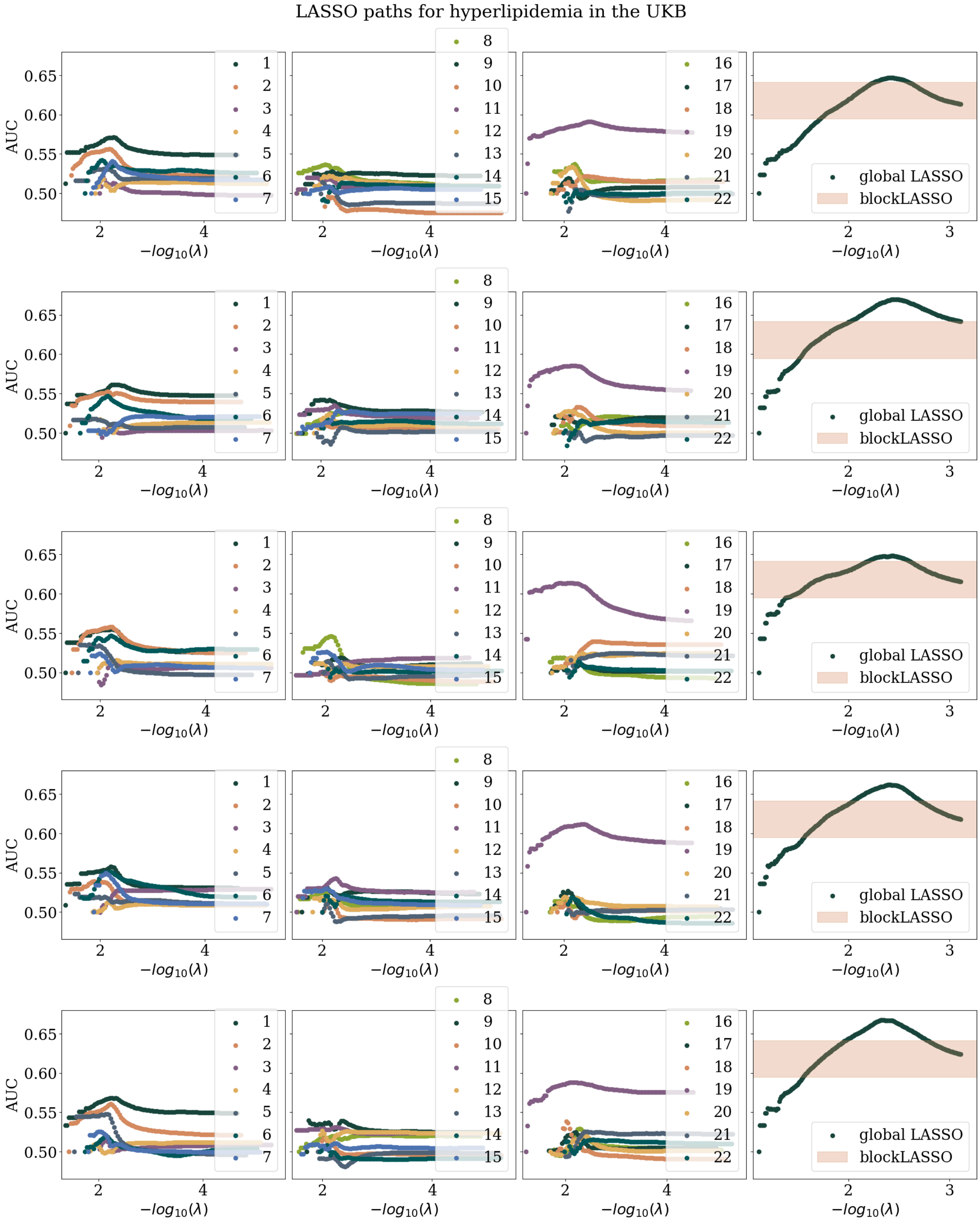

**Figure 26:** Coordinate descent based LASSO paths within the validation/model-selction set within the UKB. The first three panels left to right show the paths for each chromosome for the blockLASSO construction. The fourth panel on the right shows the comparison of the global LASSO (dots) vs the complete (i.e., re-weighted) blockLASSO value in the validation set. Different rows correspond to different cross-validation folds.

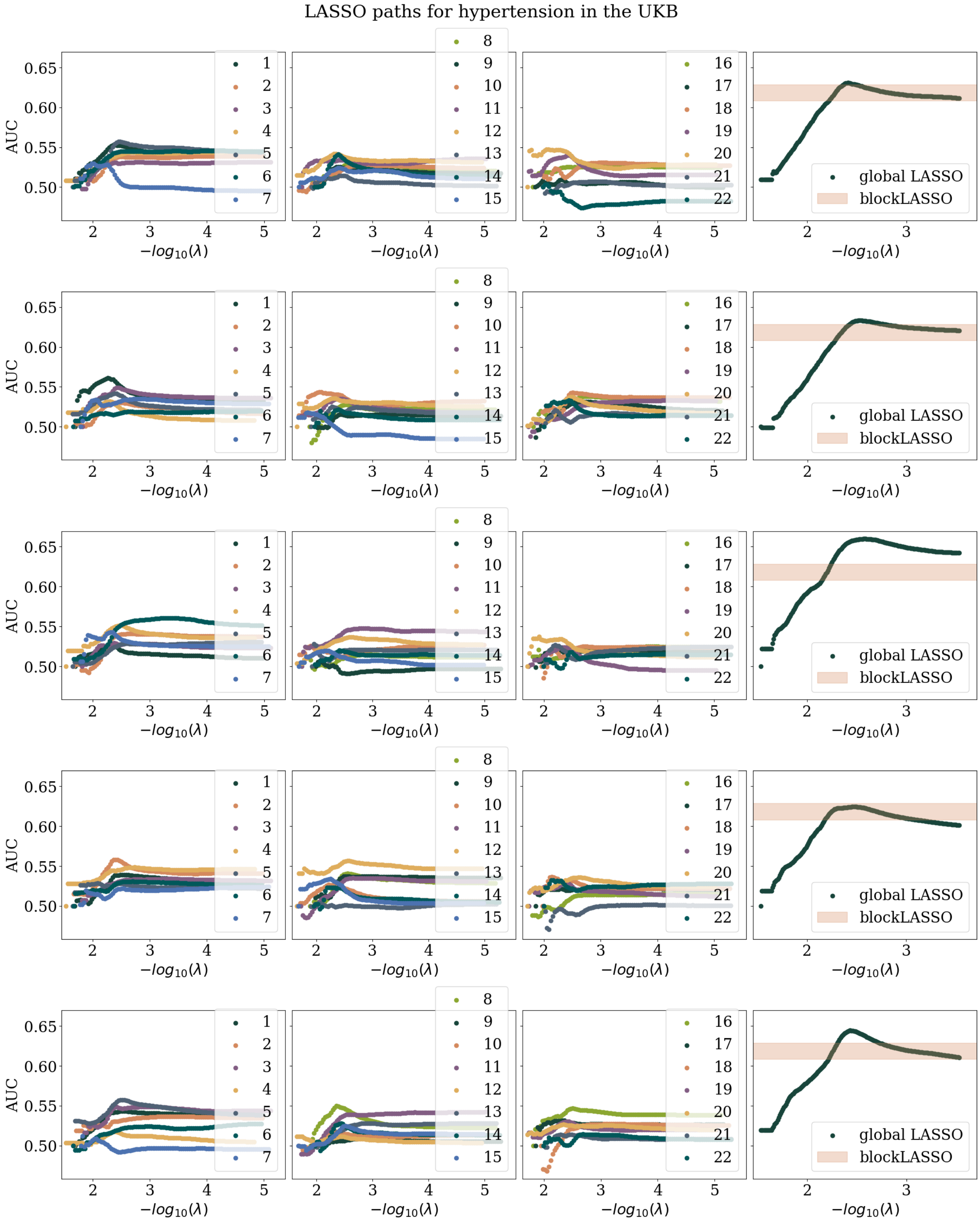

**Figure 27:** Coordinate descent based LASSO paths within the validation/model-selction set within the UKB. The first three panels left to right show the paths for each chromosome for the blockLASSO construction. The fourth panel on the right shows the comparison of the global LASSO (dots) vs the complete (i.e., re-weighted) blockLASSO value in the validation set. Different rows correspond to different cross-validation folds.

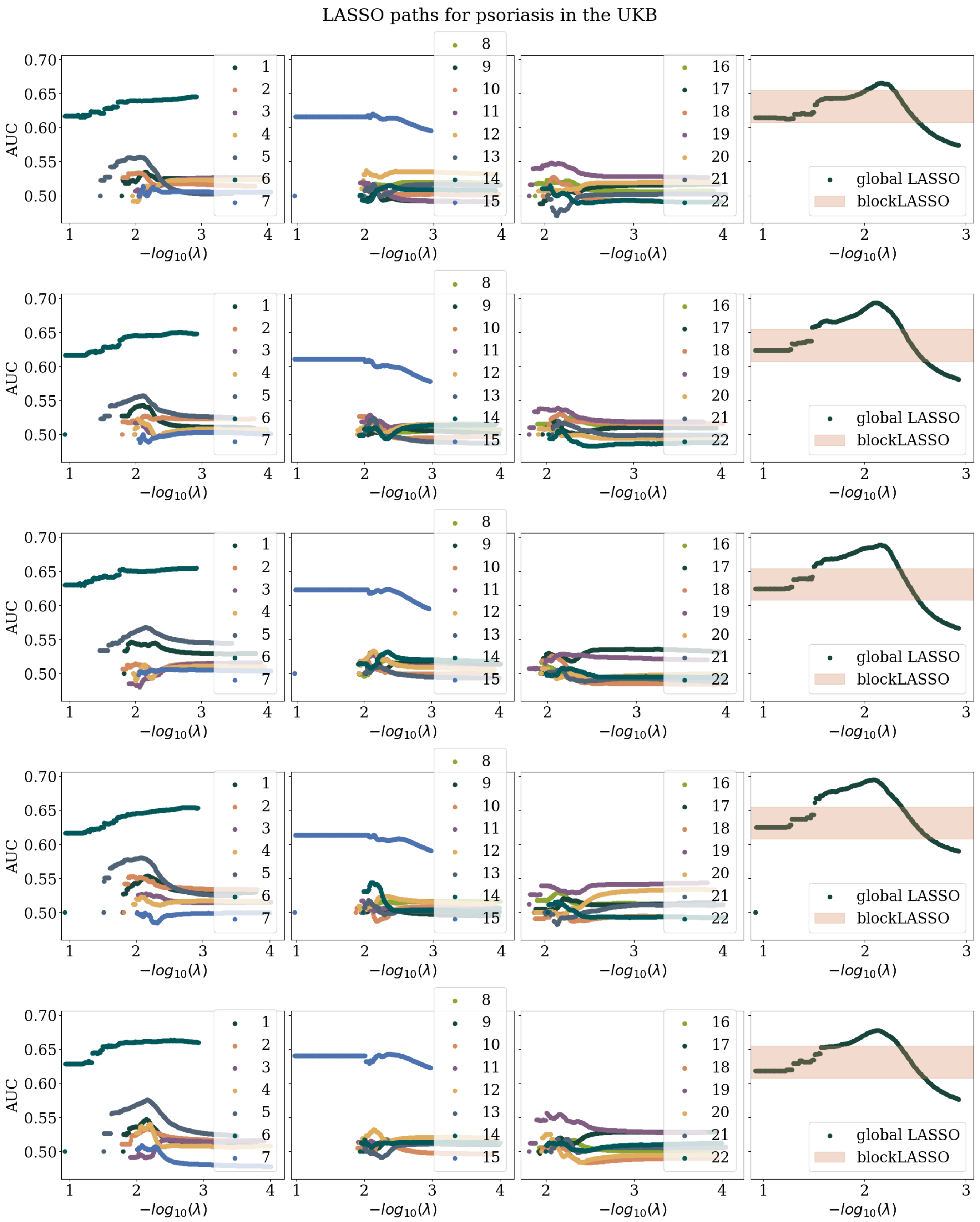

**Figure 28:** Coordinate descent based LASSO paths within the validation/model-selction set within the UKB. The first three panels left to right show the paths for each chromosome for the blockLASSO construction. The fourth panel on the right shows the comparison of the global LASSO (dots) vs the complete (i.e., re-weighted) blockLASSO value in the validation set. Different rows correspond to different cross-validation folds.

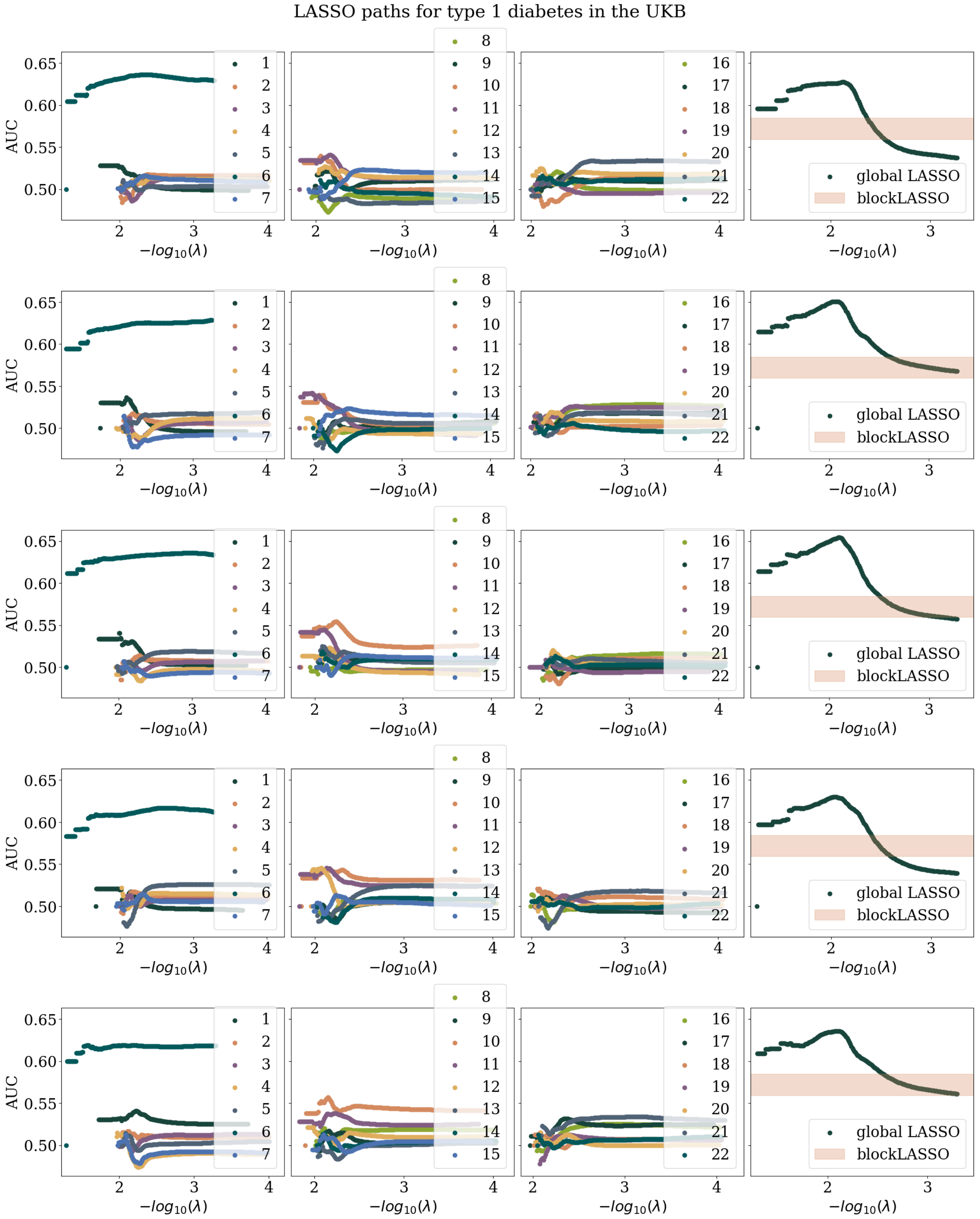

**Figure 29:** Coordinate descent based LASSO paths within the validation/model-selction set within the UKB. The first three panels left to right show the paths for each chromosome for the blockLASSO construction. The fourth panel on the right shows the comparison of the global LASSO (dots) vs the complete (i.e., re-weighted) blockLASSO value in the validation set. Different rows correspond to different cross-validation folds.

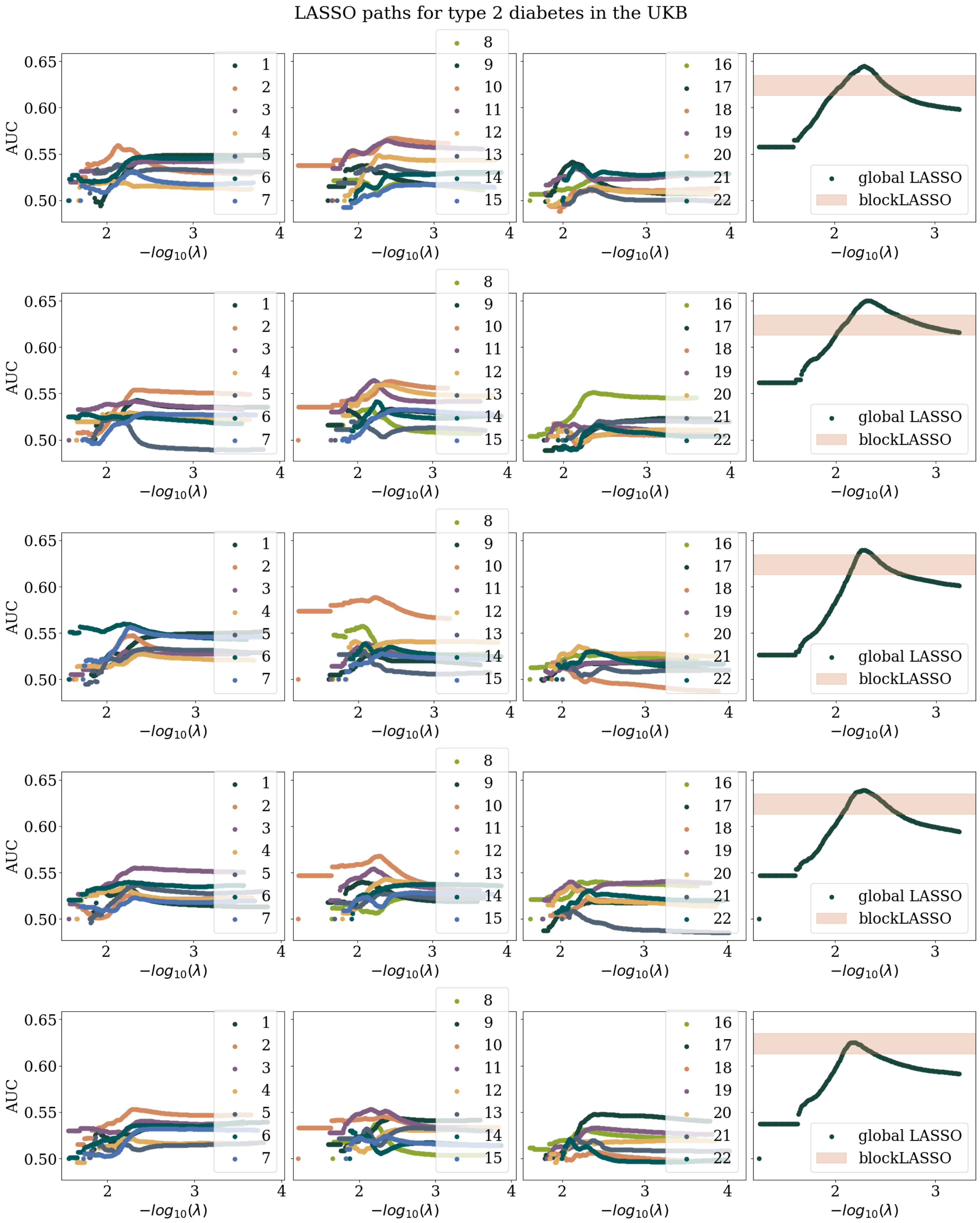

**Figure 30:** Coordinate descent based LASSO paths within the validation/model-selction set within the UKB. The first three panels left to right show the paths for each chromosome for the blockLASSO construction. The fourth panel on the right shows the comparison of the global LASSO (dots) vs the complete (i.e., re-weighted) blockLASSO value in the validation set. Different rows correspond to different cross-validation folds.

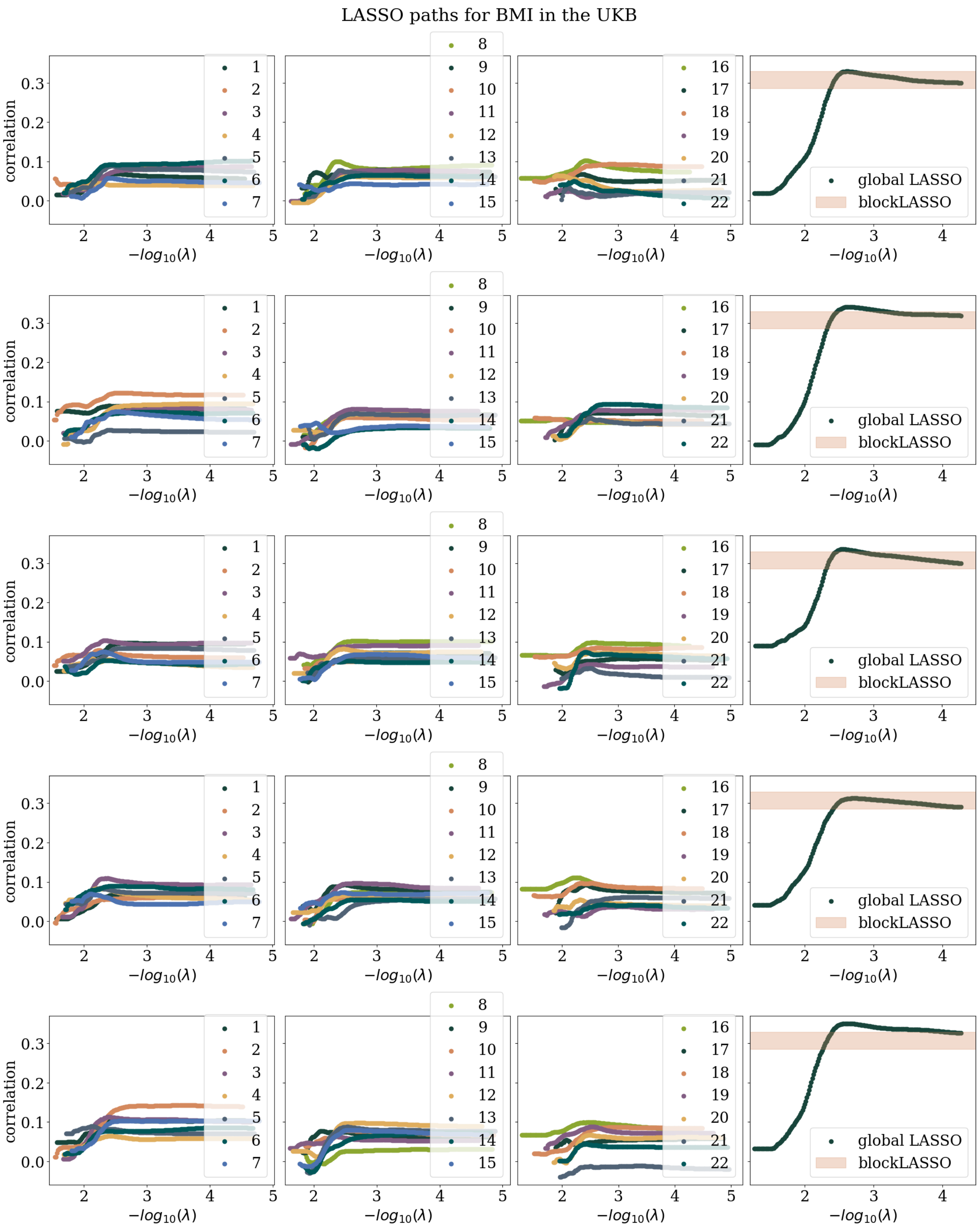

**Figure 31:** Coordinate descent based LASSO paths within the validation/model-selction set within the UKB. The first three panels left to right show the paths for each chromosome for the blockLASSO construction. The fourth panel on the right shows the comparison of the global LASSO (dots) vs the complete (i.e., re-weighted) blockLASSO value in the validation set. Different rows correspond to different cross-validation folds.

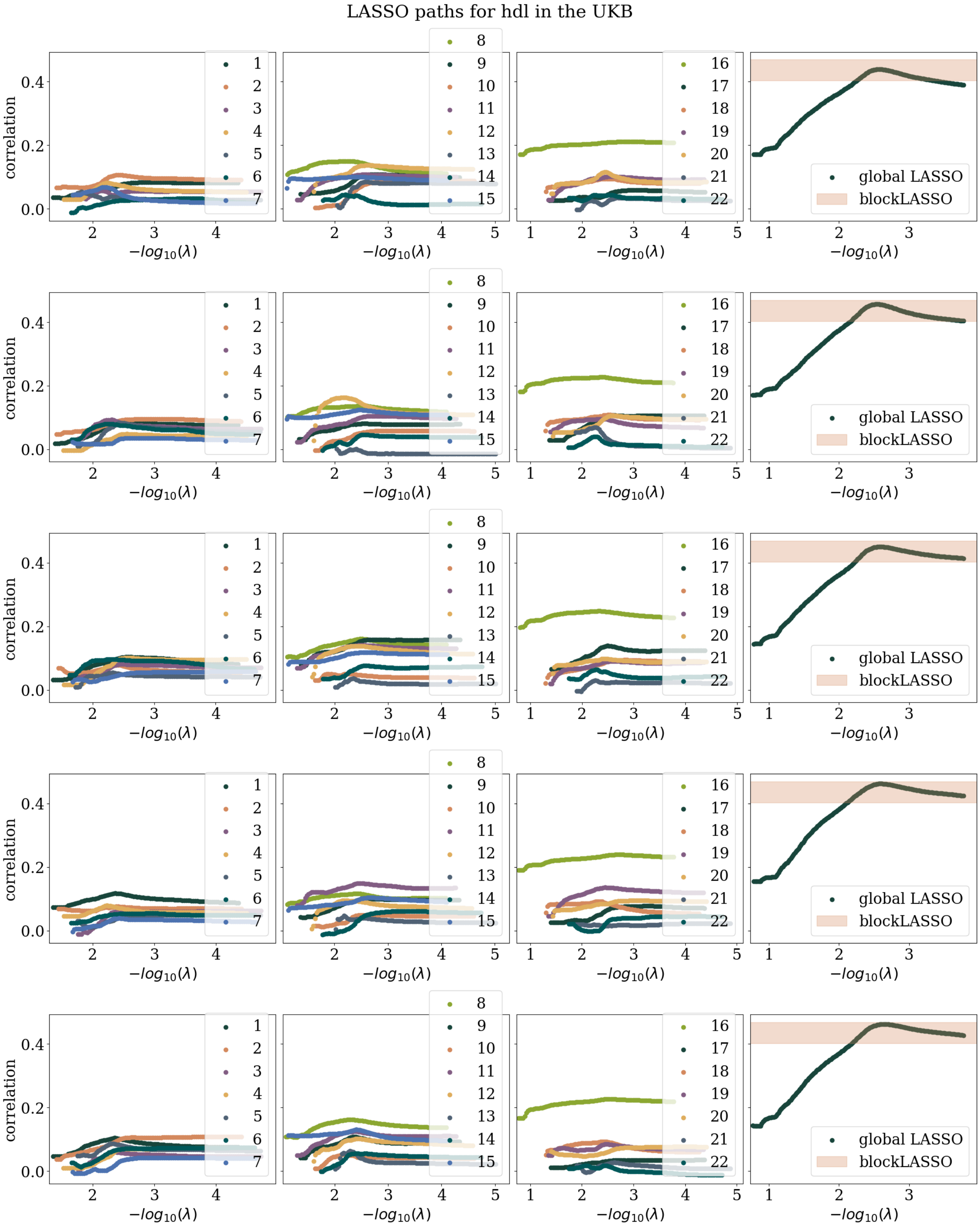

**Figure 32:** Coordinate descent based LASSO paths within the validation/model-selction set within the UKB. The first three panels left to right show the paths for each chromosome for the blockLASSO construction. The fourth panel on the right shows the comparison of the global LASSO (dots) vs the complete (i.e., re-weighted) blockLASSO value in the validation set. Different rows correspond to different cross-validation folds.

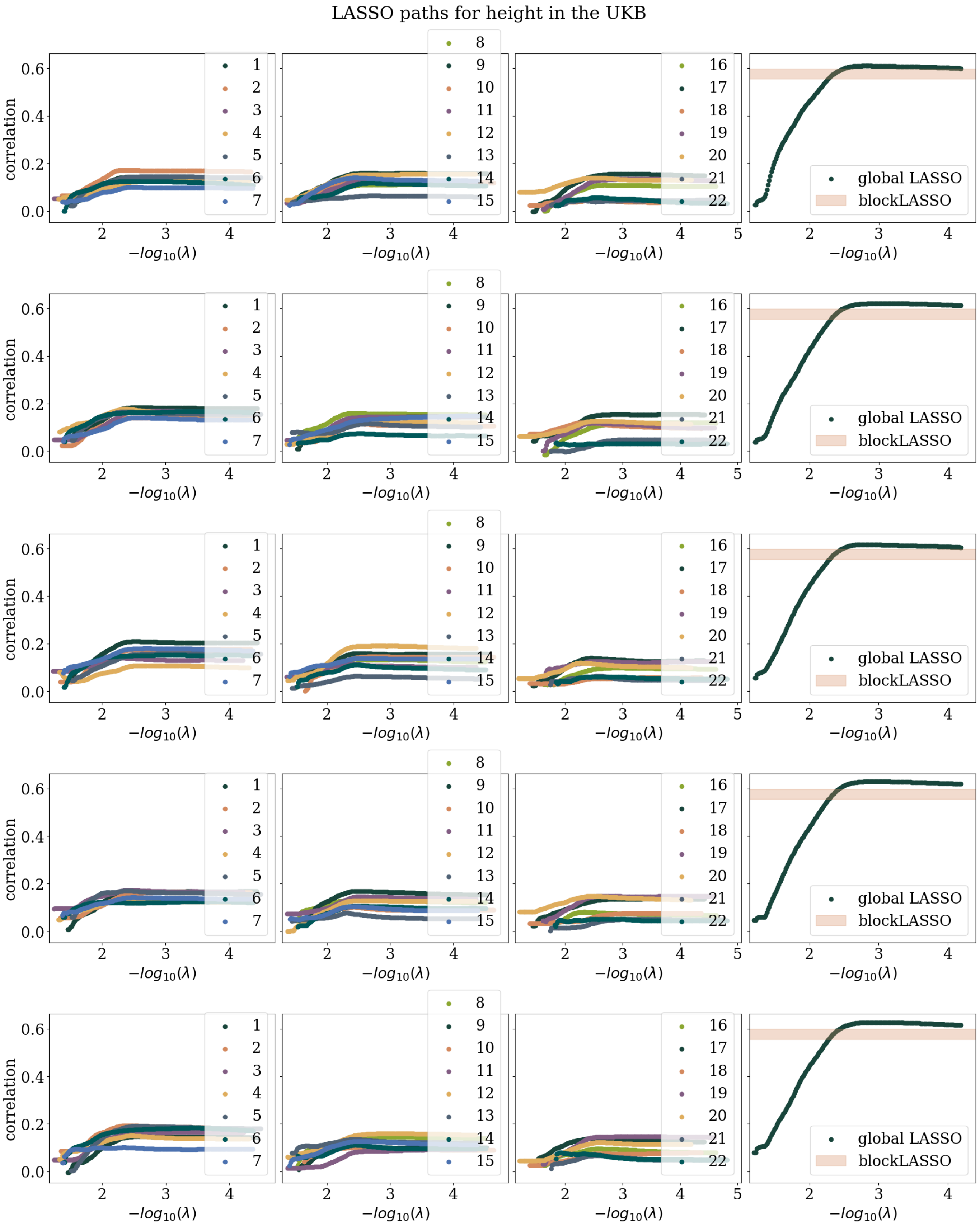

**Figure 33:** Coordinate descent based LASSO paths within the validation/model-selction set within the UKB. The first three panels left to right show the paths for each chromosome for the blockLASSO construction. The fourth panel on the right shows the comparison of the global LASSO (dots) vs the complete (i.e., re-weighted) blockLASSO value in the validation set. Different rows correspond to different cross-validation folds.

LASSO paths for total bilirubin in the UKB

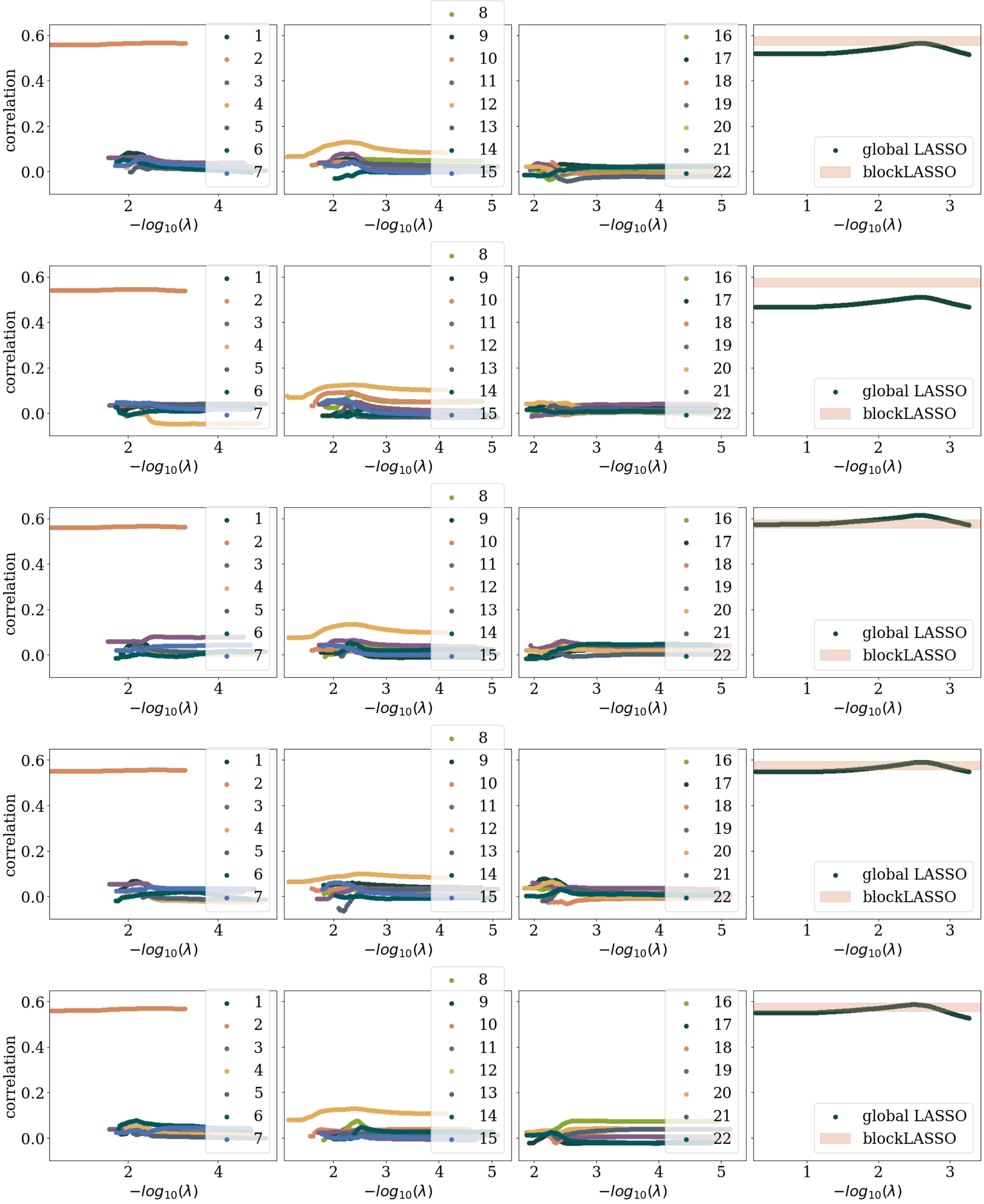

**Figure 34:** Coordinate descent based LASSO paths within the validation/model-selction set within the UKB. The first three panels left to right show the paths for each chromosome for the blockLASSO construction. The fourth panel on the right shows the comparison of the global LASSO (dots) vs the complete (i.e., re-weighted) blockLASSO value in the validation set. Different rows correspond to different cross-validation folds.

### References

1. Raben, T. G., Lello, L., Widen, E. & Hsu, S. D. Biobank-scale methods and projections for sparse polygenic prediction from machine learning. *Scientific Reports* **13,** 11662 (2023) (cit. on p. 11).
2. Jette, M. A. & Wickberg, T. *Architecture of the Slurm Workload Manager* in *Workshop on Job Scheduling Strategies for Parallel Processing* (2023), 3–23 (cit. on p. 11).
